# Richer database context improves LLM-generated EHR phenotyping algorithms

**DOI:** 10.64898/2026.10.27.26366654

**Authors:** Mengshuo Ye, Harlin Lee, Selen Gizlice, Madeline Tobias, Shawn O’Neil, Emily Pfaff, Stanley C Ahalt

**Affiliations:** UCSF Radiology and Biomedical Imaging, San Francisco, CA, USA; School of Data and Information Sciences, University of North Carolina at Chapel Hill, Chapel Hill, NC, USA; RTI International, Research Triangle Park, NC, USA; Cleveland Clinic Research, Cleveland, OH, USA; Department of Genetics, University of North Carolina at Chapel Hill School of Medicine, Chapel Hill, NC, USA; Department of Medicine, University of North Carolina at Chapel Hill School of Medicine, Chapel Hill, NC, USA

**Author notes:** Corresponding author: Harlin Lee, 211 Manning Dr, Chapel Hill, NC, 27599, USA.

**Keywords:** electronic health records, phenotyping, large language models, Observational Medical Outcomes Partnership (OMOP), SQL generation

## Abstract

**Objective:** To evaluate large language model (LLM) generation of type 2 diabetes (T2DM) phenotyping SQL and the effect of database implementation context.

**Materials and Methods:** Eight publicly accessible LLM-based systems generated SQL under progressively richer prompts using synthetic and University of North Carolina (UNC) Health electronic health record (EHR) data, both based on the Observational Medical Outcomes Partnership (OMOP) model. Outputs were compared with the Electronic Medical Records and Genomics (eMERGE) T2DM reference phenotype using SQL executability, F1, positive predictive value (PPV), and recall.

**Results:** Fewer than 55% of limited-context synthetic outputs and no limited-context UNC EHR outputs executed without modification. Detailed codebooks (L3) increased executability to 81.3% in synthetic OMOP and 100% in UNC EHR; adding sample rows (L4) increased synthetic executability to 100%. Limited-context recall was as low as 0.01, whereas high-context outputs achieved recall ≥0.99 at synthetic L4 and >0.90 at UNC L3, although UNC PPV remained substantially lower than recall. At the highest available context levels, Beta F1 ranged from 0.72–0.99 in synthetic OMOP and 0.57–0.68 in UNC EHR; all UNC models exceeded the diagnosis-code baseline (F1=0.54). The top synthetic performer fell from F1=0.99 to 0.64 on UNC EHR.

**Discussion:** Richer database context was associated with more consistent SQL executability and recall, while model rankings varied across datasets.

**Conclusion:** LLMs can generate clinically meaningful T2DM phenotyping SQL when provided with sufficient database context, but limited PPV in real-world data underscores the need for local validation.

## BACKGROUND AND SIGNIFICANCE

Electronic health record (EHR) data have become an increasingly important source of information for clinical and population health research.[1] These data are often used for phenotyping, which is the process of classifying patients into specific groups using observable characteristics found in their medical records. The results of EHR phenotyping can be used in a variety of applications, such as a screening process to identify patients who are eligible for clinical trials or to identify patients who may have a specific disease.[2] Clinician-validated phenotyping algorithms such as the Electronic Medical Records and Genomics (eMERGE) network[3] have enabled such initiatives at a large scale.

Developing executable EHR phenotyping scripts remains a time-consuming process that requires simultaneous expertise in clinical medicine, local EHR data models such as the Observational Medical Outcomes Partnership (OMOP) model, and structured query language (SQL). Unlike general text-to-SQL translation, EHR phenotyping first necessitates clinical reasoning to specify inclusion and exclusion criteria, diagnosis codes, medications, laboratory thresholds, temporal relationships, and vocabulary mappings; only after this clinical logic is established can it be translated into executable code.[4] A clinician may understand the phenotype but lack programming expertise, while an analyst who can write SQL code may lack the domain knowledge to define it correctly. This expertise gap creates a significant bottleneck, particularly at institutions without dedicated medical informatics infrastructure. This combination of clinical and technical demands makes EHR phenotyping a natural candidate task for large language models (LLMs), which have recently demonstrated strong performance in both clinical reasoning and code generation.[5,6]

LLMs have shown promise on text-to-SQL translation tasks,[7] and related work on clinical trial eligibility criteria further demonstrates that LLMs can generate OMOP-compatible SQL, though hallucination, concept-mapping errors, and execution failures remain common.[8] A key open question is how much context LLMs require to generate accurate phenotyping queries, as prior work has evaluated LLMs under fixed prompting strategies without systematically varying what context is provided.[6, 9] We address this gap by evaluating multiple LLMs on T2DM phenotyping across synthetic and real-world EHR data, using the eMERGE algorithm as a reference. Prompt levels ranged from zero-shot with no schema information, through OMOP table names and relational structure, to full-context conditions that include detailed codebooks and, where patient privacy permits, representative sample data rows.

## MATERIALS AND METHODS

### Study design

Eight LLMs were evaluated without fine-tuning: ChatGPT 5.5,[10] Claude Sonnet 4.5,[11] Gemini 3,[12] Gemma-4-31B,[13] Llama-3.3-70B,[14] Copilot (Free / Copilot Chat),[15] and two Custom GPTs designed for SQL generation (QueryGPT[16] and OMOP-SQL-Assistant,[17] both available on ChatGPT.com). They were chosen as representative, publicly accessible systems likely to be used by researchers. Longer descriptions are provided in Supplementary Material.

We prompted each LLM to generate SQL phenotyping scripts for T2DM under progressively richer database-context conditions across synthetic and real-world OMOP data. We focus on T2DM because it is widely phenotyped, has a well-validated eMERGE reference algorithm, and enables direct comparison with prior work.[6]

Model outputs were compared with the eMERGE reference phenotype using F1, positive predictive value (PPV), recall and cohort overlap. In addition, SQL executability, defined as the proportion of outputs that ran without modification, was assessed.

This study on de-identified data was determined to be Institutional Review Board (IRB) exempt by the University of North Carolina (UNC) at Chapel Hill under #25-0534.

### Datasets

The eMERGE T2DM phenotype[3] classifies cases through five mutually exclusive pathways, each requiring absence of T1DM diagnosis codes (ICD-9 250.*1, 250.*3 or ICD-10 E10.*) combined with T2DM diagnosis codes (ICD-9 250.*0, 250.*2 excluding 250.10 and 250.12, or ICD-10 E11.*), T2DM medications, or abnormal laboratory values (HbA1c ≥6.5%, fasting glucose ≥125 mg/dL, or random glucose >200 mg/dL). Path 5 additionally requires at least two physician-entered T2DM diagnoses. The algorithm was adapted to OMOP using standard concept ancestry for the synthetic dataset and source field matching for the UNC EHR dataset, with equivalent five-path logic in both implementations.

The synthetic dataset contained 124,150 total patients, of whom 6,215 were positive by the eMERGE reference phenotype. It was derived from the Synthea COVID-19 dataset[18] and manually adapted using ETL SyntheaBuilder[19] to conform to the OMOP common data model v5.3.1.

We constructed three UNC EHR patient cohorts using ICD-10 diagnosis codes recorded between October 1, 2015 and March 18, 2025, restricted to patients with at least three completed office visits or hospitalizations at UNC Health. Cohort 1 included patients with E11.* or O24.1* codes (T2DM) appearing at least three times in the patient’s problem list or the billing record. Cohort 2 included those with O24.0*, O24.3*, O24.4*, O24.8*, O24.9*, E08.*, E09.*, E10.*, or E13.* codes (other diabetes), again appearing at least three times. Cohort 3 served as a control cohort and excluded all diagnoses used to define Cohorts 1 and 2. 8,000 patients were randomly sampled from each cohort, yielding a dataset of up to 24,000 patients. Patients overlapping with Cohort 1 were excluded from Cohorts 2 and 3, prioritizing the T2DM diagnosis-based cohort assignment. Cohorts 2 and 3 retained 30 overlapping patients, who were deduplicated in the combined analytic population. The final population comprised 23,820 unique patients, including 5,732 eMERGE-positive patients. eMERGE identified 5,732 positives within this universe that served as the reference standard. Analysis was conducted within the UNC Secure Health Informatics Research Environment (SHIRE), ensuring that patient data was never transmitted to external LLMs. Data were extracted from the UNC Healthcare system on July 31, 2025, and were formatted using the OMOP Common Data Model v5.4. Therefore, although the two datasets were OMOP-based, the implementations differed in practical details including schema qualifiers, available fields, SQL dialect, and vocabulary mapping conventions.

### Prompting strategy

Prompt levels differed in the amount of database implementation context provided (Table 1). Level 0 was a zero-shot condition from an earlier set of synthetic experiments and was retained as a reference. The cross-dataset comparison began at Level 1, which provided the available OMOP table names only. Level 2 additionally described the relationships among those tables. Level 3 provided a detailed, dataset-specific codebook describing available tables, columns, vocabulary conventions, and SQL implementation details. Level 4 extended Level 3 by adding representative sample rows from the synthetic database. This condition was evaluated only on synthetic OMOP data because patient privacy restrictions prevented the inclusion of real UNC EHR records in external LLM prompts. Evaluating both Levels 3 and 4 on synthetic data allowed us to distinguish the contribution of detailed database documentation from the additional contribution of representative data examples.

**Table 1.** Prompt context levels and their application across datasets.

| Level | Context Description | Synthetic | UNC EHR |
| --- | --- | --- | --- |
| 0 | None | Yes | No |
| 1 | OMOP table names | Yes | Yes |
| 2 | OMOP table names and relations | Yes | Yes |
| 3 | Detailed codebook of schema, fields, implementation details | Yes | Yes |
| 4 | Codebook and sample rows from available tables | Yes | No, due to patient privacy constraints. |

For each context level, we generated outputs using two prompting strategies from Yan et al.[6] Alpha prompting consisted of five independent generations using the same prompt, run separately without any prior output as input (e.g., A1P1–A1P5 for Level 1), to account for response variability due to the probabilistic nature of LLMs. Beta prompting produced a single consolidated output by presenting all five Alpha outputs back to the model and asking it to assess their quality and synthesize an improved SQL script (e.g., B1 for Level 1), a strategy shown to mitigate hallucination through iterative self-refinement.[20] Detailed prompt templates are provided in the Supplementary Material.

### SQL execution and compatibility handling

For limited-context outputs (Levels 0, 1, and 2), prompts did not include schema or SQL dialect details, so when necessary, environment-specific compatibility edits were applied before evaluation. The most common OMOP-specific repair was adding source-to-standard concept mapping via the concept_relationship table (relationship_id = ‘Maps to’), so that ICD-9/ICD-10 source codes resolved to standard condition_concept_id values in condition_occurrence. These edits enabled SQL execution but did not alter the underlying phenotyping logic. In the synthetic analysis, these edits were applied to several Beta outputs when needed; Alpha outputs were left as generated to preserve direct executability as an evaluation outcome. In the UNC EHR analysis, all limited-context outputs (A1, A2, B1, B2) required compatibility edits.

## RESULTS

### Diagnosis codes alone are insufficient for reliable T2DM phenotyping

eMERGE identified 5,732 T2DM-positive patients among the 23,820 patients in the UNC EHR labeled universe (Table 2). Cohort 1, designed as the diagnosis-code-derived T2DM-positive cohort, contained only 3,708 eMERGE-positive patients out of 8,000 (PPV = 0.46, recall = 0.65, F1 = 0.54, Jaccard = 0.37). The two intended negative cohorts were also mixed, as Cohorts 2 and 3 contained 1,335 and 695 eMERGE-positive patients, respectively. This limited agreement confirms that ICD-10 diagnosis codes alone do not reliably reproduce a clinician-validated T2DM phenotype, underscoring the need for multi-step SQL algorithms of the kind evaluated in this study.

**Table 2.** Agreement between UNC EHR cohorts and the eMERGE reference phenotype.

|  | UNC EHR cohort | n | eMERGE-positive in cohort |
| --- | --- | --- | --- |
| 1 | T2DM Diagnosed | 8,000 | 3,708 |
| 2 | Other Diabetes Diagnosed | 7,884 | 1,335 |
| 3 | Control | 7,966 | 695 |

Note: Cohort sizes sum to 23,850 rather than 23,820 because 30 patients appear in both Cohorts 2 and 3 (counted once in the combined population). Six of these 30 are eMERGE-positive, so the cohort-level eMERGE-positive counts ( $3,708 + 1,335 + 695 = 5,738$ ) exceed the 5,732 unique eMERGE-positive patients.

### Full-context prompting improves SQL executability

Prompt context had a clear effect on SQL executability (Table 3). In the synthetic dataset, fewer than 55% of outputs at Levels 0–2 executed without modification. Direct executability increased to 81.3% (39/48) at Level 3 with the detailed codebook and reached 100% (48/48) at Level 4 after representative sample rows were added. In the UNC EHR setting, no Level 1 or Level 2 output executed directly, whereas all Level 3 outputs (48/48) executed without modification. These findings indicate that detailed database documentation substantially improved SQL executability, with sample-row context providing an additional improvement in the synthetic setting.

**Table 3.** Direct SQL executability by dataset and prompt context.

| Dataset | Context | Directly executable | Compatibility edits |
| --- | --- | --- | --- |
| Synthetic | Level 0 | 18 / 48 (37.5%) | 7/8 Beta outputs |
|  | Level 1 | 26 / 48 (54.2%) | 6/8 Beta outputs |
|  | Level 2 | 25 / 48 (52.1%) | 6/8 Beta outputs |
|  | Level 3 | 39 / 48 (81.3%) | 0 |
|  | Level 4 | 48 / 48 (100%) | 0 |
| UNC EHR | Level 1 | 0 / 48 (0%) | 48 |
|  | Level 2 | 0 / 48 (0%) | 48 |
|  | Level 3 | 48 / 48 (100%) | 0 |

Failure modes were heterogeneous. Non-executable outputs included references to unavailable tables or columns, invalid aliases or CTE dependencies, malformed SQL, misuse of aggregate functions, and query timeouts. Separately, some queries executed successfully but returned empty cohorts because they made incorrect assumptions about the local OMOP vocabulary implementation, including missing source-to-standard Maps to mappings or unpopulated concept_ancestor relationships. These errors indicate that limited-context models struggled with both SQL/schema alignment and local vocabulary conventions.

### Effect of prompt context on phenotyping F1

Among outputs that executed, context also improved phenotyping performance (Figure 1). In the synthetic OMOP setting (Figure 1A), low-context prompts produced highly variable results: many outputs were non-executable, and F1 scores within a single model often varied widely across the five independent Alpha attempts. For example, Claude Sonnet 4.5 Alpha outputs at Level 0 ranged from 0.10 to 0.97. Higher-context outputs (Levels 3–4) were generally more stable and achieved higher F1 scores across models than limited-context outputs (Levels 0–2), with diminishing returns beyond schema-level context for most models.

**Figure 1.**
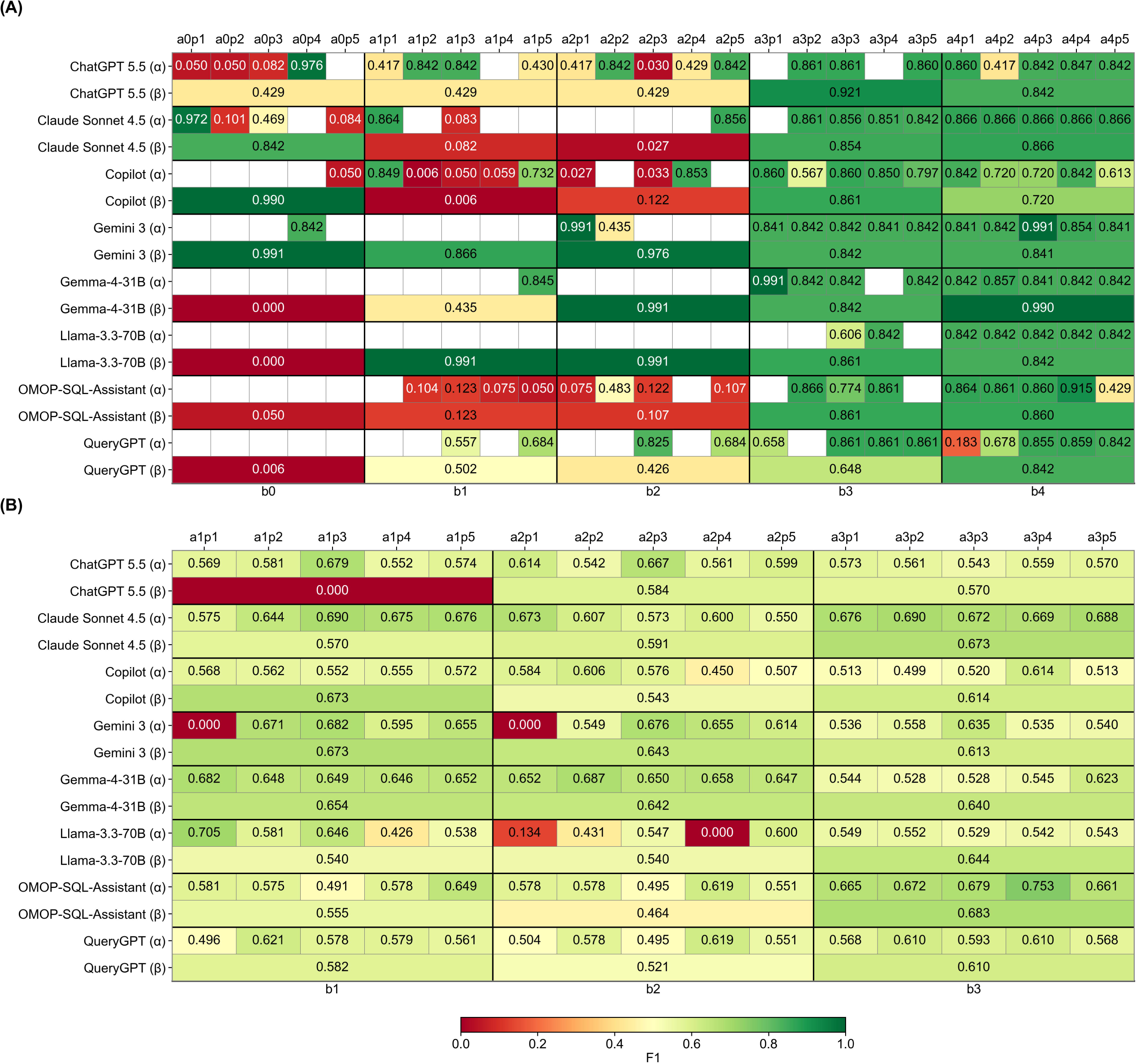
F1 heatmaps by model and prompt in (A) synthetic and (B) UNC EHR data. Rows represent each model separated by Alpha and Beta outputs, and columns represent individual prompts grouped by context level. Alpha rows contain the five independent prompt attempts within each context level, while Beta rows show the consolidated output for that same context level. Blank cells indicate outputs that did not produce an executable or evaluable SQL result. SQL compatibility edits are applied as summarized in **Table 3**. Alt text: (A) Synthetic OMOP F1 heatmap showing highly variable performance and numerous non-evaluable outputs at lower context levels, with generally higher and more consistent scores at Levels 3–4. (B) UNC EHR heatmap showing narrower F1 variation across models, with more consistent performance under full-context prompting.

In the UNC EHR setting (Figure 1B), F1 scores were more compressed across models and context levels, reflecting the greater difficulty of reproducing the eMERGE phenotype in real-world data. Nonetheless, the same pattern held: low-context outputs occasionally failed entirely (e.g., ChatGPT 5.5 Beta and Gemini Alpha at Level 1 returned F1 = 0.00), while Level 3 full-context outputs were more consistent, particularly for Beta outputs. Recall heatmaps are available in Supplementary Material.

### Context primarily increases recall, not precision

To understand what drove these F1 differences, we examined the PPV–recall decomposition of Beta outputs (Figure 2). In the synthetic dataset, low-context outputs clustered in the upper-left quadrant where PPV is high but recall is as low as 0.01, indicating that without additional database context, models generated overly restrictive queries that identified very few positive patients. Full-context outputs shifted toward the upper-right, achieving recall close to 1.0 across nearly all models at a modest cost to PPV. The same directional shift occurred in the UNC EHR setting, where full-context outputs achieved recall above 0.9 across all eight models. However, PPV ranged from 0.403 to 0.529, exceeding 0.5 for only two models, indicating that false positives remained an important limitation in real-world data.

**Figure 2.**
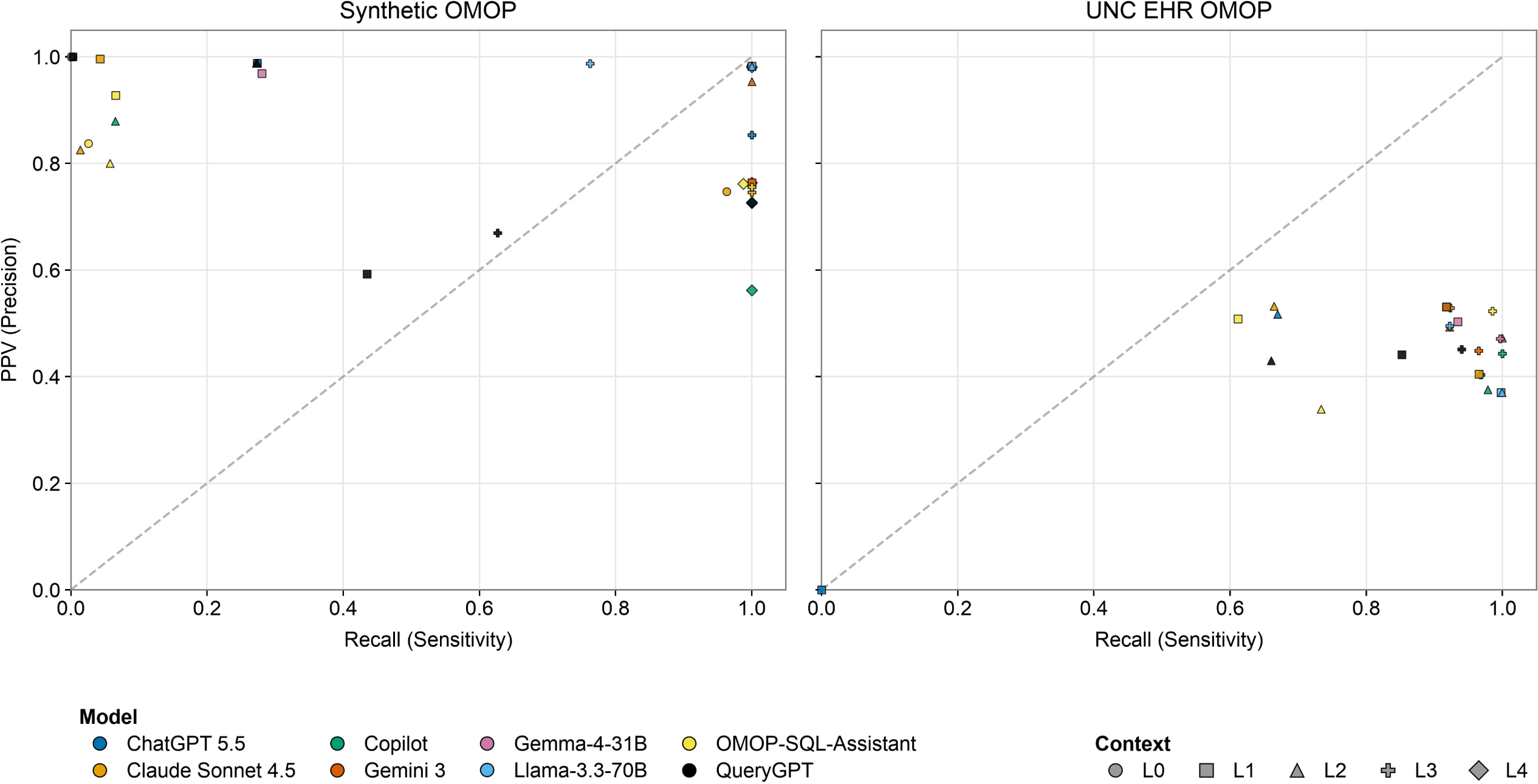
Precision-recall space for Beta outputs in synthetic (Left) and UNC EHR (Right) data. Each point represents one model at one context level, with color indicating model and marker shape indicating context. The diagonal line represents equal PPV and recall. Full-context outputs generally shifted toward higher recall in both settings, while PPV remained the main limiting dimension in the UNC EHR analysis. Alt text: (A) Synthetic OMOP precision–recall scatterplot showing that higher-context outputs generally achieve greater recall, while limited-context outputs often exhibit high precision but low recall. (B) UNC EHR scatterplot showing high recall but comparatively low precision across full-context models.

Across both settings, adding context primarily increased recall rather than precision. Without schema grounding, LLM-generated queries missed the majority of eMERGE-positive patients while maintaining high precision on the few they did capture. Full-context prompting reversed this tradeoff, favoring broader inclusion criteria.

### Higher-context queries tend toward simpler, more interpretable logic

The annotated flowcharts visualize phenotyping logic and patient counts at each decision node (Figure 3), offering a qualitative explanation for this shift. Using ChatGPT 5.5 as an illustrative example, we show that higher-context prompts tended to produce simpler and more clinically interpretable logic.

**Figure 3.**
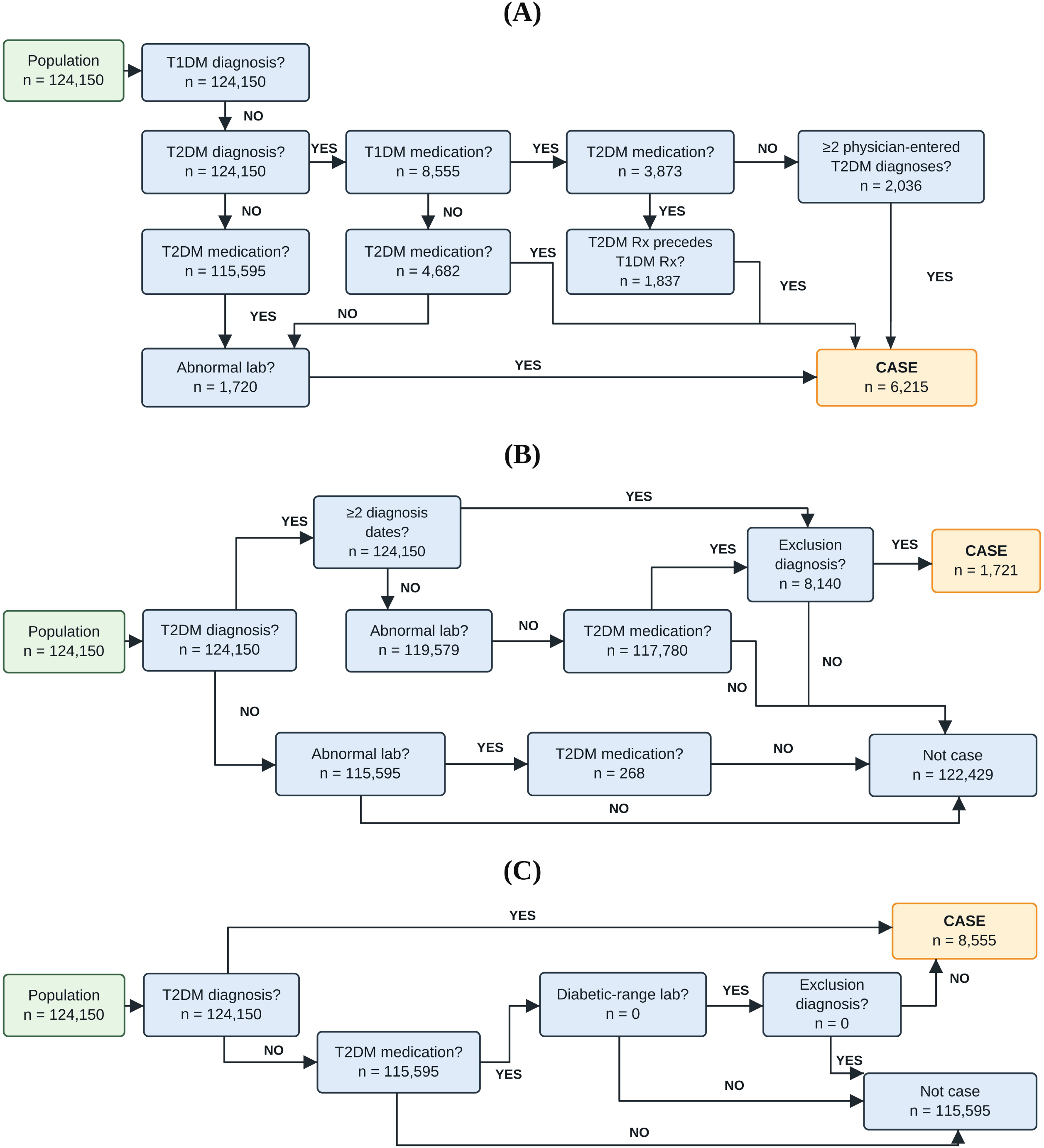
T2DM phenotyping logic flowcharts. (A) eMERGE reference adapted from PheKB with patient counts added,[21] (B) example lower-performing ChatGPT 5.5 Beta B1 flowchart (F1 = 0.429), showing a more fragmented multi-evidence rule, and (C) example higher-performing ChatGPT 5.5 Beta B4 flowchart (F1 = 0.842), showing a simpler diagnosis-first rule. Alt text: (A) Reference eMERGE phenotyping flowchart using diagnosis, medication, and laboratory criteria to identify 6,215 cases. (B) Lower-performing model flowchart with fragmented decision pathways identifying 1,721 cases. (C) Higher-context model flowchart with simpler, diagnosis-first logic identifying 8,555 cases.

The eMERGE algorithm starts from 124,150 patients and identifies 6,215 final cases (Figure 3A). The lower-performing ChatGPT 5.5 Beta B1 output (F1 = 0.429) identifies only 1,721 cases and 122,429 non-cases (Figure 3B). Its logic is fragmented across multiple alternative evidence pathways, including repeated diagnosis dates, abnormal laboratory values, medication use, and a broad downstream exclusion step. The counts show that large numbers of patients remain in intermediate branches (for example, 119,579 at one abnormal-lab node, 117,780 at a medication node, and 8,140 reaching the terminal exclusion node), helping explain why this phenotype becomes difficult to interpret and poorly aligned with the reference.

In contrast, the higher-performing ChatGPT 5.5 Beta B4 output (F1 = 0.842) identifies 8,555 cases and 115,595 non-cases (Figure 3C). Its logic is much simpler: a direct T2DM diagnosis is sufficient for case assignment, while medication plus diabetic-range laboratory evidence serves as a secondary pathway only when direct diagnosis evidence is absent. However, despite achieving perfect recall (1.000) and a higher F1 score (0.842), the B4 algorithm also identified 2,340 false positives relative to the eMERGE reference (PPV = 0.726). The pattern highlights a tradeoff: simpler, more inclusive phenotyping logic may improve case identification while introducing additional false positives. The annotated counts show that most of the classification is driven by the diagnosis-first branch, with little contribution from later exclusion filtering, making the overall logic easier to follow. This example suggests that richer database context may help models produce simpler, more interpretable phenotype definitions, although improvements in F1 do not necessarily eliminate the risk of over-inclusion.

### Beta prompting does not consistently help in limited-context settings

Having established how context shapes query logic, we examined whether the Beta prompting step added value within each context level (Table 4). Across both datasets, Beta prompting did not consistently outperform Alpha in limited-context settings. In the synthetic dataset, Beta mean F1 was lower than Alpha mean F1 at both Level 1 and Level 2. In the UNC EHR analysis, Beta mean F1 was also lower than Alpha at Level 1 and only slightly higher at Level 2. In the synthetic dataset, mean Alpha F1 decreased slightly from 0.823 at Level 3 to 0.801 at Level 4, whereas mean Beta F1 increased from 0.836 to 0.850. Beta F1 was greater than or equal to the corresponding mean Alpha F1 for six of eight models at both levels. Thus, adding representative sample rows improved aggregate Beta performance but did not consistently improve Alpha performance. In the UNC EHR setting, where Level 4 was not evaluated, Level 3 Beta outputs achieved a mean F1 of 0.631 compared with 0.588 for Alpha outputs.

**Table 4.** Alpha versus Beta performance across context levels.

| Dataset | Context | Mean Alpha F1 | Mean Beta F1 | Models where Beta F1<br>≥ Mean Alpha F1 |
| --- | --- | --- | --- | --- |
| Synthetic | Level 1 | 0.500 | 0.429 | 1/6 |
|  | Level 2 | 0.556 | 0.509 | 1/6 |
|  | Level 3 | 0.823 | 0.836 | 6/8 |
|  | Level 4 | 0.801 | 0.850 | 6/8 |
| UNC EHR | Level 1 | 0.588 | 0.531 | 3/8 |
|  | Level 2 | 0.544 | 0.566 | 2/8 |
|  | Level 3 | 0.588 | 0.631 | 6/8 |

Note: Synthetic OMOP Alpha mean F1 includes only successfully executed outputs; therefore, Levels 3 and 4 may include different sets of evaluable runs. UNC EHR Alpha results reflect performance after compatibility edits. Alpha F1 is averaged across evaluable runs and models, whereas Beta F1 is averaged across models. Comparisons between context levels are descriptive.

These results suggest that consolidation alone is not sufficient when prompts lack detailed schema and implementation context. In limited-context settings, Beta outputs may preserve or amplify errors from the initial Alpha attempts. In contrast, when full implementation context was provided, Beta prompting appeared more useful, likely because the model had enough database-specific information to consolidate candidate logic into a more coherent phenotype definition.

### No single model dominates in full-context settings

Having established that context level drives performance, we turn to whether model choice matters within full-context settings. Figure 4 compares full-context Beta F1 scores for each model across the synthetic and UNC EHR datasets. In the synthetic setting, performance was generally high: most models achieved F1 above 0.84, with Gemma-4-31B performing best (0.99) and Copilot lowest (0.72). All models exceeded the diagnosis-code-derived UNC EHR baseline (F1 = 0.54, dashed line in Figure 4).

**Figure 4.**
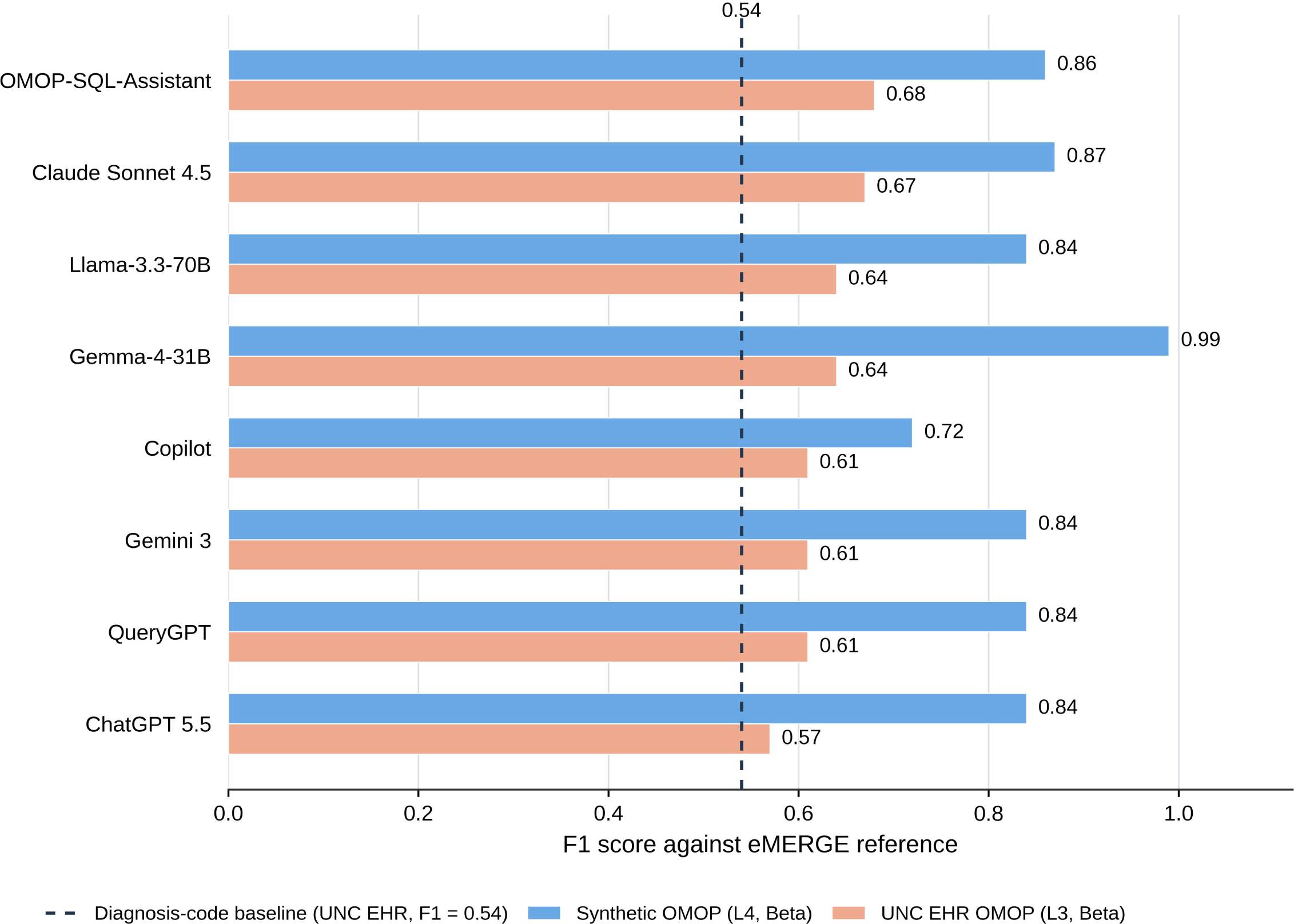
Full-context Beta F1 performance across synthetic and UNC EHR datasets. Bars show F1 at the highest available context level (Level 4 for synthetic data; Level 3 for UNC EHR). The dashed line indicates the F1 of the diagnosis-code-derived UNC EHR Cohort 1 baseline. Alt text: Horizontal bar chart comparing eight models across synthetic OMOP and UNC EHR datasets. Synthetic F1 scores range from 0.72 to 0.99, while UNC EHR scores range from 0.57 to 0.68, all exceeding the diagnosis-code baseline of 0.54.

In the more clinically relevant but difficult UNC EHR setting, performance was lower and more compressed. OMOP-SQL-Assistant and Claude Sonnet 4.5 led (F1 = 0.68 and 0.67), while ChatGPT 5.5 had the lowest F1 (0.57). The remaining five models clustered between 0.61 and 0.64. All full-context Beta outputs still exceeded the cohort-label baseline.

No model dominated across both settings. Notably, Gemma-4-31B was the top synthetic performer but ranked only mid-tier on UNC EHR data. Rankings shifted substantially between the two environments, suggesting that performance on synthetic data alone does not predict real-world accuracy in full-context settings.

## DISCUSSION

LLMs can generate executable, clinically meaningful T2DM phenotyping SQL, but performance depends critically on the amount of EHR context provided. These findings extend prior work showing that LLMs can identify relevant phenotyping concepts but produce immature logical organization without schema context,[6] and offer a practical mitigation for the hallucination and definition inconsistencies reported previously.[9] The context-dependent performance improvements observed here are also consistent with the high hallucination rates in clinical concept mapping reported in prior work,[8] suggesting that richer context may partly work by reducing concept-mapping errors.

### Context matters more than model choice or Beta prompting

The most consistent finding was that prompt context level was the primary driver of performance. In full-context settings, all eight models exceeded the diagnosis-code-derived baseline (F1=0.54), and the LLMs did not have a clear winner across environments. Even in limited-context settings, where model-intrinsic clinical knowledge might be expected to matter more, no model consistently outperformed others across datasets and context levels. Beta outputs also showed their clearest advantage over Alpha only when sufficient schema and implementation details were provided.

The incremental benefit of representative sample rows was more nuanced than that of detailed database documentation. In synthetic OMOP, adding sample rows increased direct SQL executability from 81.3% to 100%, but its effect on F1 differed between prompting strategies, with a slight decrease in mean Alpha F1 and a modest increase in mean Beta F1. These findings suggest that sample rows may improve alignment with the target database without necessarily producing more accurate phenotyping logic. Because Level 4 was not evaluated on UNC EHR data, the incremental benefit of sample-row context in real-world clinical environments remains uncertain.

The results suggest that LLMs are most useful as assistants for translating phenotype definitions into executable SQL, when prompts provide concrete schema, vocabulary, and implementation details. This reflects the dual nature of EHR phenotyping as both a semantic and an execution task. In OMOP, a correct clinical idea may still fail if the SQL uses source vocabulary codes directly when the target table stores standard concept identifiers, or if it omits required mapping steps such as the ‘Maps to’ relationship or concept_ancestor expansion. A model can generate clinically plausible logic that fails if it references nonexistent tables, unsupported functions, incompatible SQL syntax, or incomplete vocabulary mappings.

### Full-context prompting improves recall and interpretability, while PPV remains limited

Without schema grounding, models defaulted to conservative criteria, identifying few patients at high precision. Full-context prompting shifted this toward broader identification, recovering nearly all eMERGE-positive patients at the cost of more false positives. This shift was accompanied by a change in query structure: full-context outputs tended toward simpler, diagnosis-first logic, while low-context outputs produced fragmented multi-pathway rules that were both harder to interpret and less aligned with the reference phenotype. The UNC EHR full-context outputs often achieved high recall with only moderate PPV. This operating point may be useful for cohort screening, where missing true positives is costly, but it also risks broader cohorts with more false positives. The best practical phenotype may therefore depend on downstream use. For clinical trial screening, a high-recall output followed by chart review may be acceptable. For automated cohort construction without manual adjudication, higher PPV may be required.

### Performance on synthetic data does not predict real-world accuracy

The UNC EHR results show that an LLM-generated phenotype that looks strong in a clean synthetic OMOP setting may still behave differently when applied to real EHR data. Synthetic data are useful for controlled benchmarking and prompt development, but they may underestimate challenges introduced by local schemas, vocabulary conventions, SQL dialects, incomplete mappings, heterogeneous coding practices, and institution-specific cohort definitions. A practical step toward bridging this gap would be the development of standardized, shareable prompt packages, combining schema documentation, vocabulary mappings, and implementation details, that institutions could adapt for local deployment and validation. Real-world evaluation is therefore needed to assess not only whether generated phenotype logic is clinically reasonable, but also whether it remains executable, portable, and appropriately calibrated in the target data environment.

### Institutional implication for LLM-assisted phenotyping

The highest-context condition, providing sample data rows alongside schema documentation, produced the best overall performance, but schema documentation alone (Level 3) achieved comparable results for most models. This suggests that institutions can meaningfully improve LLM-assisted phenotyping through schema and codebook investment alone, without requiring direct data access, while reserving sample-row context for privacy-permitting environments such as synthetic data or secure enclaves.

### Limitations

First, eMERGE was treated as the reference standard, but no algorithm is a perfect substitute for full chart review. Second, limited-context evaluations required compatibility edits, so those results reflect phenotype logic after syntax normalization rather than zero-edit deployment. Third, models were evaluated on one disease phenotype, T2DM, and results may differ for phenotypes requiring more complex temporal logic, imaging, medications, or laboratory interpretation. Fourth, due to the cost and time required to query multiple LLMs at scale, each prompt condition was run a limited number of times, precluding formal statistical comparison of model performance differences; observed ranking differences should therefore be interpreted with caution. Fifth, the study evaluated specific model versions available at the time of testing. Because LLM capabilities, model behavior, and hosted implementations change rapidly across versions, the exact model rankings observed here may not generalize to future releases; the more durable findings may therefore be the effects of prompt context, executability, and validation strategy rather than the relative ranking of individual models.

## CONCLUSION

LLM-generated SQL phenotyping scripts can meaningfully approximate a validated T2DM algorithm when provided with sufficient database context. Full-context prompting improved SQL executability, recall, and logical interpretability across all tested models, while model choice and Beta prompting had limited influence on final performance. Synthetic benchmarking alone is insufficient to predict real-world performance, and local EHR validation remains essential. Future work should evaluate the incremental benefit of representative sample-row context in secure real-world EHR environments, assess portability and reproducibility across institutions, and incorporate clinician review of generated algorithms.

## Data Availability

Reproducibility materials, including LLM-generated SQL outputs, analysis code, and aggregate synthetic results, are publicly available at https://github.com/scottye009/llm-phenotyping. The synthetic OMOP dataset used in this study is publicly available on Zenodo (https://doi.org/10.5281/zenodo.23650498).[22] UNC EHR patient-level and row-level data cannot be shared publicly due to privacy and institutional restrictions and remain within the UNC Secure Health Informatics Research Environment (SHIRE). Full prompt templates are provided in the Supplementary Material.

## ACKNOWLEDGEMENTS

Anna Jojic and Aaron Thomas facilitated UNC EHR data acquisition. Neelasha Panda assisted with LLM prompting. Xun Zhou, Yixiang Qu, Kevin Anstrom, and Kinh Truong contributed to or gave helpful feedback to a preliminary version of this work completed as a course project at UNC. Juan Ramirez, Joyce Tan, and Jill Jemison assisted with conceptualization and project administration of a preliminary version of this study. Pratik Patekar provided assistance with navigating the SHIRE environment, and Jocelyn Tsai and Leigh Ann Samsa helped prepare the IRB application.

ChatGPT (OpenAI; GPT-6) was used to assist with manuscript language review and editorial suggestions. The authors reviewed the AI-assisted suggestions and take full responsibility for the accuracy and integrity of the final manuscript.

## AUTHOR CONTRIBUTIONS

Conceptualization: HL, EP, SCA

Data curation: MY, SO, EP

Formal analysis: MY

Funding acquisition: HL, EP, SCA

Investigation: MY

Methodology: MY, HL

Project administration: HL, EP

Resources: HL, EP, SCA

Software: MY

Supervision: HL

Validation: MY

Visualization: MY, HL

Writing – original draft: MY, HL, MT, SG

Writing – review & editing: MY, HL, MT, SG, SO, EP, SCA

