## Supplementary Material for "Richer database context improves LLM-generated EHR phenotyping algorithms"

**LLM Model Selection**

Prior work directly evaluating LLMs for EHR phenotyping SQL generation has focused on a small set of models. Yan et al.[6] evaluated GPT-4, GPT-3.5, Claude 2, and Bard (PaLM 2), the only study to date evaluating LLMs specifically for this task. Related work on phenotype definition extraction[9] and clinical trial criteria conversion to OMOP SQL[8] has similarly focused on early GPT and PaLM-generation models.

The current study extends this landscape by evaluating eight publicly accessible LLMs without fine-tuning, representing a range of model types that researchers could realistically access: updated proprietary models (ChatGPT 5.5, Claude Sonnet 4.5, Gemini 3), open and locally deployable models (Llama-3.3-70B, Gemma-4-31B), a coding-oriented assistant (Microsoft Copilot), and domain-specific SQL tools (QueryGPT, OMOP-SQL-Assistant). All models were evaluated out of the box so that results reflect practical prompt-based use rather than customized model training. This selection represents models accessible at the time of testing rather than an exhaustive ranking of LLM capability.

**ChatGPT 5.5** was included as a general-purpose OpenAI model with strong reasoning and code-generation capabilities. Because SQL phenotyping requires interpreting clinical inclusion/exclusion logic and translating it into executable database queries, ChatGPT 5.5 represents a broadly accessible general-purpose model for this task.[10]

**Claude Sonnet 4.5** was included as another widely used general-purpose proprietary model, with particular relevance for long-context reasoning and instruction following. These features are important for phenotyping prompts that include schema descriptions, variable definitions, code lists, and multi-step algorithm logic.[11]

**Gemini 3** was included as a general-purpose Google DeepMind model with multimodal and reasoning capabilities. It was selected to represent a major alternative model family that is publicly accessible and commonly used for complex reasoning and coding tasks.[12]

**Microsoft Copilot** was included because it is a widely used coding-assistant system rather than a general chatbot alone. Since the task required generating executable SQL, Copilot provided a practical comparison point for tools designed around software development and code completion.[15]

**Llama-3.3-70B** was included as an open, instruction-tuned 70-billion-parameter model. Open models are important in healthcare settings because they can potentially be deployed in controlled institutional environments, reducing reliance on external hosted systems when protected or sensitive data are involved.[14]

**Gemma-4-31B** was included as a smaller open model family designed for more accessible deployment compared with the largest proprietary systems. Including Gemma allowed us to evaluate whether a more resource-efficient open model could still generate useful SQL phenotyping logic.[13]

**OMOP-SQL-Assistant** was included because it is domain-oriented toward OMOP SQL query generation. This model represented a task-specific assistant designed around the database structure most relevant to the study, rather than a general-purpose LLM alone. Its inclusion allowed us to compare general LLMs with an OMOP-oriented SQL assistant in the same phenotyping task.[17]

**QueryGPT** was included as another domain-adapted query-generation assistant. It represented a class of tools that combine general LLM reasoning with structured query generation support, which is directly relevant to the gap between clinical phenotype definitions and executable SQL.[16]

**Preliminary model testing.** We also evaluated MedGemma during preliminary testing. Across five independent generations, only one produced a complete SQL query. The generated SQL was also not considered a reasonable T2DM phenotype. For example, it treated a T2DM diagnosis, metformin exposure, or HbA1c >6.5% as independent sufficient criteria joined by UNION, which would classify patients based on any one of these signals alone. Another output also included a Type 1 diabetes diagnosis pathway as a positive branch. Given the low rate of complete SQL generation and the limited clinical coherence of the generated logic, MedGemma was not included in the final model comparison.

An earlier round of testing used ChatGPT 5.0. When ChatGPT 5.5 became available, we replaced the earlier model with the newer version and reran the evaluation. Therefore, all ChatGPT results reported in the final analysis correspond to ChatGPT 5.5.

**Prompting**

Complete prompt templates for each context level are provided below. Dataset-specific implementation instructions were retained as used during evaluation, including PostgreSQL and representative sample-row context for synthetic Level 4 and DuckDB/SHIRE codebook context for UNC EHR Level 3.

**Alpha - L0**

You are an expert in disease phenotyping.

Please provide the phenotyping algorithm for type 2 diabetes.

Let's think step by step:

1. List the critical criteria to consider.

2. Determine how these criteria should be combined.

3. Derive the final algorithm.

Instructions to follow:

1. Use "AND", "OR", and "NOT" to organize the algorithm.

2. Use Observational Medical Outcomes Partnership (OMOP) concepts for diagnoses, symptoms, procedures, laboratory tests, medication, etc.

3. Use ICD9CM and ICD10CM codes for diagnosis concepts.

4. Use both generic names and brand names of medications for medication concepts.

5. Use SQL style to organize the final algorithm.

**Beta - L0**

You are an expert in disease phenotyping.

Below are five SQL queries (in triple quotation marks) for phenotyping type 2 diabetes in an EHR database. Your task is to check the quality of these algorithms and provide a refined version so that the cohort of type 2 diabetes can be accurately found from an EHR database that follows the OMOP Common Data Model.

Let us think step by step.

1. Read each of the five algorithms, summarize the correct criteria, specify the missing criteria, if any, and specify the incorrect criteria, if any.

2. Considering the summarization, generate a refined version of the phenotyping algorithm as the final algorithm.

Instructions to follow when generating the final algorithm:

1. Use ICD9CM and ICD10CM codes for diagnosis concepts.

2. Use the OMOP concept names for the concepts that are not diagnoses.

3. Use both generic names and brand names of medications for medication concepts.

4. Generate a single SQL query and do not use the WITH clause.

5. Do not create placeholders for any concepts.

Query 1: “““”””

Query 2: “““”””

Query 3: “““”””

Query 4: “““”””

Query 5: “““”””

**Alpha - L1**

You are an expert in disease phenotyping. Please provide the phenotyping algorithm for type 2 diabetes. Let's think step by step: 1. List the critical criteria to consider. 2. Determine how these criteria should be combined. 3. Derive the final algorithm. Instructions to follow: 1. Use "AND", "OR", and "NOT" to organize the algorithm. 2. Use Observational Medical Outcomes Partnership (OMOP) concepts for diagnoses, symptoms, procedures, laboratory tests, medication, etc. 3. Use ICD9CM and ICD10CM codes for diagnosis concepts. 4. Use both generic names and brand names of medications for medication concepts. 5. Use SQL style to organize the final algorithm. Output the SQL code, adjust based on these table names, and know that the schema is public.

person

observation_period

visit_occurrence

visit_detail

condition_occurrence

drug_exposure

procedure_occurrence

device_exposure

measurement

observation

note

note_nlp

specimen

fact_relationship

survey_conduct

location

location_history

care_site

provider

payer_plan_period

cost

drug_era

dose_era

condition_era

metadata

cdm_source

concept

vocabulary

domain

concept_class

concept_relationship

relationship

concept_synonym

concept_ancestor

source_to_concept_map

drug_strength

cohort

cohort_definition

**Beta - L1**

You are an expert in disease phenotyping. Below are five SQL queries (in triple quotation marks) for phenotyping type 2 diabetes in an EHR database. Your task is to check the quality of these algorithms and provide a refined version so that the cohort of type 2 diabetes can be accurately found from an EHR database that follows the OMOP Common Data Model. Let us think step by step.

1. Read each of the five algorithms, summarize the correct criteria, specify the missing criteria, if any, and specify the incorrect criteria, if any.

2. Considering the summarization, generate a refined version of the phenotyping algorithm as the final algorithm.

Instructions to follow when generating the final algorithm: 1. Use ICD9CM and ICD10CM codes for diagnosis concepts. 2. Use the OMOP concept names for the concepts that are not diagnoses. 3. Use both generic names and brand names of medications for medication concepts. 4. Generate a single SQL query. 5. Do not create placeholders for any concepts.

Query 1: “““”””

Query 2: “““”””

Query 3: “““”””

Query 4: “““”””

Query 5: “““”””

person

observation_period

visit_occurrence

visit_detail

condition_occurrence

drug_exposure

procedure_occurrence

device_exposure

measurement

observation

note

note_nlp

specimen

fact_relationship

survey_conduct

location

location_history

care_site

provider

payer_plan_period

cost

drug_era

dose_era

condition_era

metadata

cdm_source

concept

vocabulary

domain

concept_class

concept_relationship

relationship

concept_synonym

concept_ancestor

source_to_concept_map

drug_strength

cohort

cohort_definition

**Alpha - L2**

You are an expert in disease phenotyping. Please provide the phenotyping algorithm for type 2 diabetes. Let's think step by step: 1. List the critical criteria to consider. 2. Determine how these criteria should be combined. 3. Derive the final algorithm. Instructions to follow: 1. Use "AND", "OR", and "NOT" to organize the algorithm. 2. Use Observational Medical Outcomes Partnership (OMOP) concepts for diagnoses, symptoms, procedures, laboratory tests, medication, etc. 3. Use ICD9CM and ICD10CM codes for diagnosis concepts. 4. Use both generic names and brand names of medications for medication concepts. 5. Use SQL style to organize the final algorithm. Output the SQL code, adjust based on this image, and know that the schema is public.

OMOP CDM

Standardized clinical data:
 Person
 Observation_period
 Death
 Visit_occurrence
 Visit_detail
 Condition_occurrence
 Drug_exposure
 Procedure_occurrence
 Device_exposure
 Measurement
 Observation
 Note
 Note_NLP
 Episode
 Episode_event
 Specimen
 Fact_relationship

Standardized health system:
 Location
 Care_site
 Provider

Standardized vocabularies:
 Concept
 Vocabulary
 Domain
 Concept_class
 Concept_synonym
 Concept_relationship
 Relationship
 Concept_ancestor
 Source_to_concept_map
 Drug_strength

Standardized health economics:
 Cost
 Payer_plan_period

Standardized derived elements:
 Condition_era
 Drug_era
 Dose_era

Results schema:
 Cohort
 Cohort_definition

Standardized metadata:
 CDM_source
 Metadata

Main relationships:
 Person connects to Observation_period, Death, Visit_occurrence, Condition_occurrence, Drug_exposure, Procedure_occurrence, Device_exposure, Measurement, Observation, Note, Episode, and Specimen.
 Visit_occurrence connects to Visit_detail, Condition_occurrence, Drug_exposure, Procedure_occurrence, Device_exposure, Measurement, Observation, and Note.
 Note connects to Note_NLP.
 Episode connects to Episode_event.
 Specimen connects to Fact_relationship.
 Clinical event tables connect to standardized vocabularies, with Concept as the main hub.
 The vocabulary area connects to Condition_era, Drug_era, and Dose_era.
 Cohort_definition connects to Cohort.

**Beta - L2**

You are an expert in disease phenotyping. Below are five SQL queries (in triple quotation marks) for phenotyping type 2 diabetes in an EHR database. Your task is to check the quality of these algorithms and provide a refined version so that the cohort of type 2 diabetes can be accurately found from an EHR database that follows the OMOP Common Data Model. Let us think step by step.

1. Read each of the five algorithms, summarize the correct criteria, specify the missing criteria, if any, and specify the incorrect criteria, if any.

2. Considering the summarization, generate a refined version of the phenotyping algorithm as the final algorithm.

Instructions to follow when generating the final algorithm: 1. Use ICD9CM and ICD10CM codes for diagnosis concepts. 2. Use the OMOP concept names for the concepts that are not diagnoses. 3. Use both generic names and brand names of medications for medication concepts. 4. Generate a single SQL query. 5. Do not create placeholders for any concepts.

Query 1: “““”””

Query 2: “““”””

Query 3: “““”””

Query 4: “““”””

Query 5: “““”””

OMOP CDM

Standardized clinical data:
 Person
 Observation_period
 Death
 Visit_occurrence
 Visit_detail
 Condition_occurrence
 Drug_exposure
 Procedure_occurrence
 Device_exposure
 Measurement
 Observation
 Note
 Note_NLP
 Episode
 Episode_event
 Specimen
 Fact_relationship

Standardized health system:
 Location
 Care_site
 Provider

Standardized vocabularies:
 Concept
 Vocabulary
 Domain
 Concept_class
 Concept_synonym
 Concept_relationship
 Relationship
 Concept_ancestor
 Source_to_concept_map
 Drug_strength

Standardized health economics:
 Cost
 Payer_plan_period

Standardized derived elements:
 Condition_era
 Drug_era
 Dose_era

Results schema:
 Cohort
 Cohort_definition

Standardized metadata:
 CDM_source
 Metadata

Main relationships:
 Person connects to Observation_period, Death, Visit_occurrence, Condition_occurrence, Drug_exposure, Procedure_occurrence, Device_exposure, Measurement, Observation, Note, Episode, and Specimen.
 Visit_occurrence connects to Visit_detail, Condition_occurrence, Drug_exposure, Procedure_occurrence, Device_exposure, Measurement, Observation, and Note.
 Note connects to Note_NLP.
 Episode connects to Episode_event.
 Specimen connects to Fact_relationship.
 Clinical event tables connect to standardized vocabularies, with Concept as the main hub.
 The vocabulary area connects to Condition_era, Drug_era, and Dose_era.
 Cohort_definition connects to Cohort.

**Alpha - L3**

You are an expert in disease phenotyping. Please provide the phenotyping algorithm for type 2 diabetes. Let’s think step by step:

1. List the critical criteria to consider.

2. Determine how these criteria should be combined.

3. Derive the final algorithm.

Instructions:

1. Generate directly executable SQL.

2. Use the codebook below to choose tables and columns.

3. Prefer source-value/source-name fields for diagnoses, labs, medications, procedures, and observations.

4. Use ICD9CM and ICD10CM source-code patterns for diagnosis concepts.

5. Use both generic names and brand names of medications for medication concepts.

6. Use measurement source fields and numeric values for laboratory criteria.

7. Return person_id.

8. Add concise SQL comments.

*[UNC EHR (SHIRE) codebook / Synthetic codebook]*

Disease: Type 2 diabetes mellitus.

Please provide a final directly executable SQL query.

**Beta - L3**

You are an expert in disease phenotyping and SQL generation.

Below are five SQL queries in triple quotation marks for phenotyping type 2 diabetes in an EHR database. Your task is to check the quality of these algorithms and provide a refined version so that the cohort of type 2 diabetes can be accurately found from the database.

Before reviewing the five SQL queries, read the codebook below. This database is OMOP-like, but standardized vocabulary mapping might not be fully reliable. When judging the candidate SQL, consider whether it uses columns that are available and reliable. Prefer source-value/source-name fields when generating the final refined SQL.

*[UNC EHR (SHIRE) codebook / Synthetic codebook]*

Now review the five SQL algorithms below:

Query 1: “““”””

Query 2: “““”””

Query 3: “““”””

Query 4: “““”””

Query 5: “““”””

Task:

Check the quality of these algorithms and provide a refined version so that the cohort of type 2 diabetes can be accurately found.

**Alpha - L4**

You are an expert in disease phenotyping and SQL generation.

Please provide the phenotyping algorithm for type 2 diabetes in a synthetic OMOP CDM database. Let’s think step by step:

1. List the critical criteria to consider.

2. Determine how these criteria should be combined.

3. Derive the final algorithm.

Database setup:

- Use PostgreSQL SQL for pgAdmin4.

- Use the public schema, for example public.condition_occurrence.

- Do not use DuckDB-only syntax.

- Do not use REGEXP_LIKE; use PostgreSQL regex operators such as ~ or ~*.

- Use only tables and columns shown in the sample rows below.

OMOP implementation notes:

- This synthetic OMOP database has populated clinical concept fields and vocabulary tables.

- Available vocabulary tables include concept, concept_relationship, concept_ancestor, vocabulary, and relationship.

- Clinical event tables use concept IDs such as condition_concept_id, drug_concept_id, and measurement_concept_id.

- For ICD9CM/ICD10CM diagnosis logic, consider mapping source ICD concepts through concept_relationship where relationship_id = 'Maps to', then compare the mapped concept_id_2 to condition_occurrence.condition_concept_id.

- Do not assume ICD concept_id values can be directly compared to condition_concept_id.

- For medications and labs, use drug_concept_id and measurement_concept_id when populated; use source fields only if needed.

Available tables and sample rows:

*Table Examples*

Disease:

Type 2 diabetes mellitus.

Instructions:

1. Use ICD9CM and ICD10CM concepts for diagnosis logic.

2. Use both generic and brand names for medication concepts.

3. Use OMOP concept names or concept codes for lab concepts when possible.

4. Include appropriate exclusion logic if supported by the available tables.

5. Generate one directly executable PostgreSQL SQL query.

6. Return person_id.

7. Add concise SQL comments.

8. Do not invent unavailable columns or placeholders.

Please provide the final directly executable PostgreSQL SQL query.

**Beta - L4**

You are an expert in disease phenotyping and SQL generation.

Below are five SQL queries in triple quotation marks for phenotyping type 2 diabetes in a synthetic OMOP CDM database. Your task is to review them and provide one refined SQL query that accurately identifies the T2DM cohort.

Database setup:

- Use PostgreSQL SQL for pgAdmin4.

- Use the public schema, for example public.condition_occurrence.

- Do not use DuckDB-only syntax.

- Do not use REGEXP_LIKE; use PostgreSQL regex operators such as ~ or ~*.

- Use only tables and columns shown in the sample rows below.

OMOP implementation notes:

- This synthetic OMOP database has populated clinical concept fields and vocabulary tables.

- Available vocabulary tables include concept, concept_relationship, concept_ancestor, vocabulary, and relationship.

- Clinical event tables use concept IDs such as condition_concept_id, drug_concept_id, and measurement_concept_id.

- For ICD9CM/ICD10CM diagnosis logic, check whether candidate SQL incorrectly compares ICD concept IDs directly to condition_concept_id.

- When needed, map ICD source concepts through concept_relationship where relationship_id = 'Maps to', then compare concept_id_2 to condition_occurrence.condition_concept_id.

- For medications and labs, prefer drug_concept_id and measurement_concept_id when populated; use source fields only if needed.

Available tables and sample rows:

*Table Examples*

Query 1: “““”””

Query 2: “““”””

Query 3: “““”””

Query 4: “““”””

Query 5: “““”””

Task:

1. Briefly review each algorithm.

2. Identify correct, missing, and incorrect criteria.

3. Identify SQL patterns that may fail in PostgreSQL or this OMOP implementation.

4. Generate one refined final SQL query.

Final SQL instructions:

1. Generate a single directly executable PostgreSQL SQL query.

2. Use public.table_name references.

3. Use only tables and columns shown in the sample rows.

4. Use ICD9CM and ICD10CM concepts for diagnosis logic.

5. Use concept_relationship with relationship_id = 'Maps to' when mapping ICD source diagnosis concepts to standard condition concepts is needed.

6. Use both generic and brand medication names.

7. Use OMOP concept names/codes for medication and lab concepts when possible.

8. Do not create placeholders.

9. Return person_id.

10. Add concise SQL comments.

Response format: Final refined PostgreSQL SQL query.

**Synthetic codebook**

Synthetic OMOP database codebook for SQL generation

Query engine:

- Use PostgreSQL SQL (executed through pgAdmin 4).

- Tables are in the public schema (e.g., public.person).

- Use ILIKE for case-insensitive matching and ~ or ~* for regular expressions.

- Use CAST(column AS DATE) or column::date for date casting.

- Use PostgreSQL INTERVAL syntax for date arithmetic.

- Do not reference CSV files, DuckDB views, or DuckDB-only functions.

- Use the exact column names documented below; do not invent unavailable fields.

General implementation notes:

- This synthetic database follows an OMOP-style schema with populated standard concept IDs and vocabulary tables.

- person_id is the patient join key; visit_occurrence_id and visit_detail_id link clinical events to encounters.

- Clinical *_concept_id fields can join concept.concept_id to retrieve names and codes; 0 indicates missing/unmapped concepts.

- For ICD9CM/ICD10CM codes, use concept and available concept_relationship mappings (e.g., 'Maps to') to resolve standardized concepts. Diagnosis source values may use SNOMED codes rather than ICD codes.

- concept_ancestor supports hierarchy expansion where mappings are populated; verify mapping coverage.

- measurement_source_value commonly contains LOINC codes; drug_source_value may contain numeric drug codes. Clinical event tables do not contain *_concept_name columns.

- Use value_as_number together with unit_concept_id or unit_source_value for numeric lab thresholds; account for missing/zero fields.

- These codebook notes describe schema and implementation, not a disease-specific phenotype definition.

Tables with documented columns:

- person, condition_occurrence, drug_exposure, measurement, observation, visit_occurrence, visit_detail, procedure_occurrence, death, concept, concept_relationship, concept_ancestor, vocabulary, relationship

Other available tables (column details are not provided here):

- all_visits, assign_all_visit_ids, care_site, cdm_source, concept_class, concept_synonym, condition_era, device_exposure, domain, drug_era, drug_strength, fake_claims_data, fake_patient_data, final_visit_ids, provider, source_to_standard_vocab_map

person:

- Columns:

- person_id, gender_concept_id, year_of_birth, month_of_birth, day_of_birth, birth_datetime, race_concept_id, ethnicity_concept_id

- location_id, provider_id, care_site_id, person_source_value, gender_source_value, gender_source_concept_id, race_source_value, race_source_concept_id

- ethnicity_source_value, ethnicity_source_concept_id

- Field reliability notes:

- person_id is the patient join key; birth_datetime and year_of_birth support age calculations.

- Some demographic source_concept_id fields and ethnicity_concept_id may be 0.

condition_occurrence:

- Columns:

- condition_occurrence_id, person_id, condition_concept_id, condition_start_date, condition_start_datetime, condition_end_date, condition_end_datetime, condition_type_concept_id

- condition_status_concept_id, stop_reason, provider_id, visit_occurrence_id, visit_detail_id, condition_source_value, condition_source_concept_id, condition_status_source_value

- Field reliability notes:

- condition_concept_id is the standardized diagnosis; condition_source_value often contains a SNOMED code.

- Source and standard concept IDs may differ; condition_status_concept_id may be 0.

drug_exposure:

- Columns:

- drug_exposure_id, person_id, drug_concept_id, drug_exposure_start_date, drug_exposure_start_datetime, drug_exposure_end_date, drug_exposure_end_datetime, verbatim_end_date

- drug_type_concept_id, stop_reason, refills, quantity, days_supply, sig, route_concept_id, lot_number

- provider_id, visit_occurrence_id, visit_detail_id, drug_source_value, drug_source_concept_id, route_source_value, dose_unit_source_value

- Field reliability notes:

- drug_concept_id is the standardized drug ID; drug_source_value is commonly a numeric code, not a drug name.

- Join concept for drug names; use drug_strength for ingredient mapping when available. Some dose/administrative fields are 0 or blank.

measurement:

- Columns:

- measurement_id, person_id, measurement_concept_id, measurement_date, measurement_datetime, measurement_time, measurement_type_concept_id, operator_concept_id

- value_as_number, value_as_concept_id, unit_concept_id, range_low, range_high, provider_id, visit_occurrence_id, visit_detail_id

- measurement_source_value, measurement_source_concept_id, unit_source_value, value_source_value

- Field reliability notes:

- measurement_concept_id identifies the test; measurement_source_value often contains a LOINC code.

- Use value_as_number for thresholds and unit_source_value or unit_concept_id for units; some unit and value concept IDs are 0.

observation:

- Columns:

- observation_id, person_id, observation_concept_id, observation_date, observation_datetime, observation_type_concept_id, value_as_number, value_as_string

- value_as_concept_id, qualifier_concept_id, unit_concept_id, provider_id, visit_occurrence_id, visit_detail_id, observation_source_value, observation_source_concept_id

- unit_source_value, qualifier_source_value

- Field reliability notes:

- observation_concept_id identifies the observation; observation_source_value may contain SNOMED or LOINC codes.

- Value fields may be empty and concept fields may be 0; there is no value_source_value column.

visit_occurrence:

- Columns:

- visit_occurrence_id, person_id, visit_concept_id, visit_start_date, visit_start_datetime, visit_end_date, visit_end_datetime, visit_type_concept_id

- provider_id, care_site_id, visit_source_value, visit_source_concept_id, admitting_source_concept_id, admitting_source_value, discharge_to_concept_id, discharge_to_source_value

- preceding_visit_occurrence_id

- Field reliability notes:

- visit_occurrence_id links clinical events to encounters; visit_concept_id supports visit-type classification.

- visit_source_value may be an encounter identifier rather than descriptive text.

visit_detail:

- Columns:

- visit_detail_id, person_id, visit_detail_concept_id, visit_detail_start_date, visit_detail_start_datetime, visit_detail_end_date, visit_detail_end_datetime, visit_detail_type_concept_id

- provider_id, care_site_id, visit_detail_source_value, visit_detail_source_concept_id, admitting_source_value, admitting_source_concept_id, discharge_to_source_value, discharge_to_concept_id

- preceding_visit_detail_id, visit_detail_parent_id, visit_occurrence_id

- Field reliability notes:

- visit_detail_id links granular encounter records; visit_occurrence_id links to visit_occurrence.

- visit_detail_source_value may be an encounter identifier; use the exact listed admitting/discharge column names.

procedure_occurrence:

- Columns:

- procedure_occurrence_id, person_id, procedure_concept_id, procedure_date, procedure_datetime, procedure_type_concept_id, modifier_concept_id, quantity

- provider_id, visit_occurrence_id, visit_detail_id, procedure_source_value, procedure_source_concept_id, modifier_source_value

- Field reliability notes:

- procedure_concept_id and procedure_source_value identify procedures; source and standard concept IDs may differ.

- provider_id and quantity may be blank.

death:

- Columns:

- person_id, death_date, death_datetime, death_type_concept_id, cause_concept_id, cause_source_value, cause_source_concept_id

- Field reliability notes:

- death_date and death_datetime are present; cause_concept_id and cause_source_value identify cause when populated.

concept:

- Columns:

- concept_id, concept_name, domain_id, vocabulary_id, concept_class_id, standard_concept, concept_code, valid_start_date

- valid_end_date, invalid_reason

- Field reliability notes:

- concept_id joins vocabularies to clinical records; concept_name and concept_code provide readable terms/codes.

- standard_concept marks standard concepts (commonly S); concept_id 0 means No matching concept.

concept_relationship:

- Columns:

- concept_id_1, concept_id_2, relationship_id, valid_start_date, valid_end_date, invalid_reason

- Field reliability notes:

- concept_id_1, concept_id_2, and relationship_id identify mapping direction; Maps to links source to standard concepts where present.

- Check invalid_reason and validity dates when mapping.

concept_ancestor:

- Columns:

- ancestor_concept_id, descendant_concept_id, min_levels_of_separation, max_levels_of_separation

- Field reliability notes:

- ancestor_concept_id to descendant_concept_id supports hierarchy expansion; separation 0 may represent self.

- Not every vocabulary concept is covered by the same hierarchy.

vocabulary:

- Columns:

- vocabulary_id, vocabulary_name, vocabulary_reference, vocabulary_version, vocabulary_concept_id

- Field reliability notes:

- Lists available coding systems, including ICD9CM, ICD10CM, SNOMED, LOINC, RxNorm, and UCUM.

relationship:

- Columns:

- relationship_id, relationship_name, is_hierarchical, defines_ancestry, reverse_relationship_id, relationship_concept_id

- Field reliability notes:

- Describes relationship types; actual concept-to-concept links are in concept_relationship.

**UNC EHR (SHIRE) codebook**

SHIRE database codebook for SQL generation

Query engine:

- Use DuckDB SQL.

- Tables are available as DuckDB views.

- Use ILIKE for case-insensitive text matching.

- Do not mention files or CSV paths.

- Do not use PostgreSQL-only syntax such as ::date.

- Use CAST(date_column AS DATE) when date casting is needed.

- Do not use ILIKE ANY, LIKE ANY, = ANY, <> ALL, or other ANY/ALL comparison patterns.

- For multiple text patterns, write explicit OR conditions.

- If many keywords are needed, create an inline VALUES table and use EXISTS.

- Use regexp_matches(string, pattern) for regex matching when needed.

General implementation notes:

- The database is OMOP-like, but standardized vocabulary fields are inconsistently populated.

- Many *_concept_id fields are 0.

- Many *_concept_name fields are “No matching concept”.

- Source-value and source-name fields are usually more informative than standardized vocabulary mappings.

- The concept, concept_relationship, concept_ancestor, source_to_concept_map, LOINC, RxNorm, SNOMED, and Maps to workflow are not available/reliable for this SHIRE setup.

- All views include cohort and filename columns created by the DuckDB runner.

Available views:

- person

- condition_occurrence

- drug_exposure

- measurement

- observation

- procedure_occurrence

- visit_occurrence

- visit_detail

- death

Not currently available as DuckDB views:

- care_site

- provider

person:

- Columns:

- cohort

- person_id

- gender_concept_id

- gender_concept_name

- year_of_birth

- month_of_birth

- day_of_birth

- birth_datetime

- race_concept_id

- race_concept_name

- ethnicity_concept_id

- ethnicity_concept_name

- location_id

- provider_id

- care_site_id

- person_source_value

- gender_source_value

- gender_source_concept_id

- race_source_value

- race_source_concept_id

- ethnicity_source_value

- ethnicity_source_concept_id

- filename

- Field reliability notes:

- person_id is the main patient join key.

- year_of_birth and birth_datetime are available for age logic.

- gender/race/ethnicity source fields are available but may be encoded.

condition_occurrence:

- Columns:

- cohort

- condition_occurrence_id

- person_id

- condition_concept_id

- condition_concept_name

- condition_start_date

- condition_start_datetime

- condition_end_date

- condition_end_datetime

- condition_type_concept_id

- condition_type_concept_name

- stop_reason

- provider_id

- visit_occurrence_id

- visit_detail_id

- condition_source_value

- condition_source_concept_id

- condition_source_concept_vocabulary_id

- condition_source_concept_name

- condition_status_source_value

- condition_status_concept_id

- condition_status_concept_name

- filename

- Field reliability notes:

- condition_source_value is the main field for diagnosis source codes.

- condition_source_concept_vocabulary_id is available and may identify the source vocabulary.

- condition_source_concept_name is available and may contain human-readable diagnosis text.

- condition_concept_id and condition_concept_name may be populated for some rows, but source fields are more reliable for phenotype matching.

- condition_status_concept_id and condition_status_concept_name may be 0 or “No matching concept”.

drug_exposure:

- Columns:

- cohort

- drug_exposure_id

- person_id

- drug_concept_id

- drug_concept_name

- drug_exposure_start_date

- drug_exposure_start_datetime

- drug_exposure_end_date

- drug_exposure_end_datetime

- verbatim_end_date

- drug_type_concept_id

- drug_type_concept_name

- stop_reason

- refills

- quantity

- days_supply

- sig

- route_concept_id

- route_concept_name

- lot_number

- provider_id

- visit_occurrence_id

- visit_detail_id

- drug_source_value

- drug_source_concept_id

- drug_source_concept_code

- route_source_value

- dose_unit_source_value

- filename

- Field reliability notes:

- drug_source_value, drug_concept_name, drug_source_concept_code, and drug_type_concept_name are available medication-related fields.

- drug_source_concept_name is not available.

- drug_concept_id and drug_source_concept_id may be populated, but RxNorm hierarchy/mapping should not be assumed.

measurement:

- Columns:

- cohort

- measurement_id

- person_id

- measurement_concept_id

- measurement_concept_name

- measurement_date

- measurement_datetime

- measurement_time

- measurement_type_concept_id

- measurement_type_concept_name

- operator_concept_id

- operator_concept_name

- value_as_number

- value_as_concept_id

- value_as_concept_name

- unit_concept_id

- unit_concept_name

- range_low

- range_high

- provider_id

- visit_occurrence_id

- visit_detail_id

- measurement_source_value

- measurement_source_concept_id

- unit_source_value

- value_source_value

- unit_source_concept_id

- measurement_event_id

- meas_event_field_concept_id

- filename

- Field reliability notes:

- measurement_source_value is the main field for identifying lab/test names.

- value_as_number is available for numeric lab thresholds.

- unit_source_value and unit_concept_name are available for unit logic.

- measurement_concept_id may be 0 and measurement_concept_name may be “No matching concept”.

- value_as_concept_id and value_as_concept_name may be 0 or “No matching concept”.

- LOINC concept IDs should not be assumed usable.

observation:

- Columns:

- cohort

- observation_id

- person_id

- observation_concept_id

- observation_concept_name

- observation_date

- observation_datetime

- observation_type_concept_id

- observation_type_concept_name

- value_as_number

- value_as_string

- value_as_concept_id

- value_as_concept_name

- qualifier_concept_id

- qualifier_concept_name

- unit_concept_id

- unit_concept_name

- provider_id

- visit_occurrence_id

- visit_detail_id

- observation_source_value

- observation_source_concept_id

- unit_source_value

- qualifier_source_value

- value_source_value

- observation_event_id

- obs_event_field_concept_id

- filename

- Field reliability notes:

- observation_source_value, value_as_string, and value_source_value may contain clinically useful text.

- observation_concept_id and observation_concept_name may be populated for broad categories.

- Many value/unit concept fields may be 0 or “No matching concept”.

procedure_occurrence:

- Columns:

- cohort

- procedure_occurrence_id

- person_id

- procedure_concept_id

- procedure_concept_name

- procedure_date

- procedure_datetime

- procedure_type_concept_id

- procedure_type_concept_name

- modifier_concept_id

- modifier_concept_name

- quantity

- provider_id

- visit_occurrence_id

- visit_detail_id

- procedure_source_value

- procedure_source_concept_id

- modifier_source_value

- procedure_end_date

- procedure_end_datetime

- filename

- Field reliability notes:

- procedure_source_value is available for procedure matching.

- procedure_source_concept_name is not available.

- procedure_concept_id and procedure_source_concept_id may be populated for some rows, but standard procedure vocabulary mapping should not be assumed.

visit_occurrence:

- Columns:

- cohort

- person_id

- visit_occurrence_id

- visit_concept_id

- visit_concept_name

- visit_start_date

- visit_start_datetime

- visit_end_date

- visit_end_datetime

- visit_type_concept_id

- visit_type_concept_name

- provider_id

- care_site_id

- visit_source_value

- visit_source_concept_id

- admitted_from_concept_id

- admitted_from_source_value

- discharged_to_concept_id

- discharged_to_source_value

- preceding_visit_occurrence_id

- filename

- Field reliability notes:

- visit_concept_name and visit_source_value are available for visit-type logic.

- visit_occurrence_id can link visits to condition_occurrence, measurement, drug_exposure, procedure_occurrence, observation, and visit_detail.

- admitted/discharged concept fields may be 0; source values may be more informative.

visit_detail:

- Columns:

- cohort

- visit_detail_id

- person_id

- visit_detail_concept_id

- visit_detail_concept_name

- visit_detail_start_date

- visit_detail_start_datetime

- visit_detail_end_date

- visit_detail_end_datetime

- visit_detail_type_concept_id

- visit_detail_type_concept_name

- provider_id

- care_site_id

- admitted_from_concept_id

- discharged_to_concept_id

- preceding_visit_detail_id

- visit_detail_source_value

- visit_detail_source_concept_id

- admitted_from_source_value

- discharged_to_source_value

- parent_visit_detail_id

- visit_occurrence_id

- filename

- Field reliability notes:

- visit_detail_id is available.

- visit_occurrence_id links visit_detail back to visit_occurrence.

- visit_detail_concept_name and visit_detail_source_value are available for granular visit-type logic.

- visit_detail_source_concept_id may be 0.

death:

- Columns:

- cohort

- person_id

- death_date

- death_datetime

- death_type_concept_id

- death_type_concept_name

- cause_concept_id

- cause_source_value

- cause_source_concept_id

- filename

- Field reliability notes:

- death_date and death_datetime are available.

- cause fields may be sparsely populated or unavailable for phenotype logic.

**Table Examples**

Selected table samples:

The following sample rows are from selected tables most relevant to phenotype SQL generation, including clinical event tables and vocabulary/mapping tables. They are included only to show table structure, column names, value formats, and whether standard/source fields are populated.

The selected tables shown are:

person, condition_occurrence, drug_exposure, measurement, observation, visit_occurrence, visit_detail, procedure_occurrence, death, concept, concept_relationship, concept_ancestor, vocabulary, relationship.

The database also contains 16 additional tables that are not shown here:

all_visits, assign_all_visit_ids, care_site, cdm_source, concept_class, concept_synonym, condition_era, device_exposure, domain, drug_era, drug_strength, fake_claims_data, fake_patient_data, final_visit_ids, provider, source_to_standard_vocab_map.

person

"person_id"	"gender_concept_id"	"year_of_birth"	"month_of_birth"	"day_of_birth"	"birth_datetime"	"race_concept_id"	"ethnicity_concept_id"	"location_id"	"provider_id"	"care_site_id"	"person_source_value"	"gender_source_value"	"gender_source_concept_id"	"race_source_value"	"race_source_concept_id"	"ethnicity_source_value"	"ethnicity_source_concept_id"

1	8532	1958	12	2	"1958-12-02 00:00:00"	8527	0	"0001049f-9248-47fe-b479-ea80eb51ce4a"	"F"	0	"white"	0	"nonhispanic"	0

2	8532	1945	10	2	"1945-10-02 00:00:00"	8527	0	"0001054e-77c4-4e09-8310-7e5b1a579d8a"	"F"	0	"white"	0	"nonhispanic"	0

3	8507	1968	4	20	"1968-04-20 00:00:00"	8527	0	"00019dce-e502-4272-82d2-d1c280f74150"	"M"	0	"white"	0	"nonhispanic"	0

4	8507	1914	7	28	"1914-07-28 00:00:00"	8527	0	"0001ae39-67c6-443c-81e4-e835ab4f6a9b"	"M"	0	"white"	0	"hispanic"	0

5	8532	1988	8	9	"1988-08-09 00:00:00"	8527	0	"0002760d-9404-43cb-873f-26b0a59d3a55"	"F"	0	"white"	0	"nonhispanic"	0

6	8532	1976	8	9	"1976-08-09 00:00:00"	8527	0	"00034773-ca66-4191-8f43-56a9c20f8abf"	"F"	0	"white"	0	"nonhispanic"	0

7	8532	1958	7	25	"1958-07-25 00:00:00"	8516	0	"00037885-87aa-4086-aa89-f56ddf5d30e0"	"F"	0	"black"	0	"nonhispanic"	0

8	8532	2017	3	27	"2017-03-27 00:00:00"	8527	0	"00039f5a-cfff-4bd4-9a0b-1768734ec059"	"F"	0	"white"	0	"nonhispanic"	0

9	8507	1972	12	22	"1972-12-22 00:00:00"	8527	0	"00050455-3247-450f-899e-709079405df8"	"M"	0	"white"	0	"nonhispanic"	0

10	8532	1998	10	3	"1998-10-03 00:00:00"	8527	0	"00051dc6-38dc-4781-a6db-b69479990236"	"F"	0	"white"	0	"nonhispanic"	0

condition_occurrence

"condition_occurrence_id"	"person_id"	"condition_concept_id"	"condition_start_date"	"condition_start_datetime"	"condition_end_date"	"condition_end_datetime"	"condition_type_concept_id"	"condition_status_concept_id"	"stop_reason"	"provider_id"	"visit_occurrence_id"	"visit_detail_id"	"condition_source_value"	"condition_source_concept_id"	"condition_status_source_value"

160994	20342	4289517	"2020-03-11"	"2020-03-11 00:00:00"	"2020-04-03"	"2020-04-03 00:00:00"	38000175	0	514680	1514680	"36955009"	4289517

160995	20342	37311061	"2020-03-11"	"2020-03-11 00:00:00"	"2020-04-03"	"2020-04-03 00:00:00"	38000175	0	514680	1514680	"840539006"	37311061

160996	20342	442752	"2020-03-11"	"2020-03-11 00:00:00"	"2020-04-03"	"2020-04-03 00:00:00"	38000175	0	514680	1514680	"68962001"	442752

160997	20342	4223659	"2020-03-11"	"2020-03-11 00:00:00"	"2020-04-03"	"2020-04-03 00:00:00"	38000175	0	514680	1514680	"84229001"	4223659

160998	20342	4089228	"2020-03-11"	"2020-03-11 00:00:00"	"2020-04-03"	"2020-04-03 00:00:00"	38000175	0	514680	1514680	"248595008"	4089228

160999	20342	254761	"2020-03-11"	"2020-03-11 00:00:00"	"2020-04-03"	"2020-04-03 00:00:00"	38000175	0	514680	1514680	"49727002"	254761

161000	20343	437663	"2020-03-09"	"2020-03-09 00:00:00"	"2020-04-07"	"2020-04-07 00:00:00"	38000175	0	514689	1514689	"386661006"	437663

161001	20343	37311061	"2020-03-09"	"2020-03-09 00:00:00"	"2020-04-07"	"2020-04-07 00:00:00"	38000175	0	514689	1514689	"840539006"	37311061

161002	20343	4147326	"2020-03-09"	"2020-03-09 00:00:00"	"2020-04-07"	"2020-04-07 00:00:00"	38000175	0	514689	1514689	"267102003"	4147326

161003	20343	4051466	"2006-02-18"	"2006-02-18 00:00:00"	38000175	0	514685	1514685	"233678006"	4051466

drug_exposure

"drug_exposure_id"	"person_id"	"drug_concept_id"	"drug_exposure_start_date"	"drug_exposure_start_datetime"	"drug_exposure_end_date"	"drug_exposure_end_datetime"	"verbatim_end_date"	"drug_type_concept_id"	"stop_reason"	"refills"	"quantity"	"days_supply"	"sig"	"route_concept_id"	"lot_number"	"provider_id"	"visit_occurrence_id"	"visit_detail_id"	"drug_source_value"	"drug_source_concept_id"	"route_source_value"	"dose_unit_source_value"

2076099	61070	1539463	"2014-09-23"	"2014-09-23 00:00:00"	"2015-09-23"	"2015-09-23 00:00:00"	"2015-09-23"	38000175	0	0	365	0	0	0	1541374	2541374	"314231"	1539463

2076100	61070	19018935	"2009-02-18"	"2009-02-18 00:00:00"	"2010-02-24"	"2010-02-24 00:00:00"	"2010-02-24"	38000175	0	0	371	0	0	0	1541458	2541458	"197604"	19018935

2076101	61070	40169281	"1988-10-26"	"1988-10-26 00:00:00"	"1989-11-01"	"1989-11-01 00:00:00"	"1989-11-01"	38000175	0	0	371	0	0	0	1541438	2541438	"896209"	40169281

2076102	61070	19106768	"2009-02-18"	"2009-02-18 00:00:00"	"2010-02-24"	"2010-02-24 00:00:00"	"2010-02-24"	38000175	0	0	371	0	0	0	1541458	2541458	"429503"	19106768

2076103	61070	40169281	"2009-02-18"	"2009-02-18 00:00:00"	"2010-02-24"	"2010-02-24 00:00:00"	"2010-02-24"	38000175	0	0	371	0	0	0	1541458	2541458	"896209"	40169281

2076104	61070	40171902	"2009-02-18"	"2009-02-18 00:00:00"	"2010-02-24"	"2010-02-24 00:00:00"	"2010-02-24"	38000175	0	0	371	0	0	0	1541458	2541458	"897718"	40171902

2076105	61070	40163554	"2009-02-18"	"2009-02-18 00:00:00"	"2010-02-24"	"2010-02-24 00:00:00"	"2010-02-24"	38000175	0	0	371	0	0	0	1541458	2541458	"855332"	40163554

2076106	61070	19106768	"1988-10-26"	"1988-10-26 00:00:00"	"1989-11-01"	"1989-11-01 00:00:00"	"1989-11-01"	38000175	0	0	371	0	0	0	1541438	2541438	"429503"	19106768

2076107	61070	19106768	"1938-01-12"	"1938-01-12 00:00:00"	"1939-01-18"	"1939-01-18 00:00:00"	"1939-01-18"	38000175	0	0	371	0	0	0	1541391	2541391	"429503"	19106768

2076108	61070	1539463	"1985-09-30"	"1985-09-30 00:00:00"	"1986-09-30"	"1986-09-30 00:00:00"	"1986-09-30"	38000175	0	0	365	0	0	0	1541341	2541341	"314231"	1539463

measurement

"measurement_id"	"person_id"	"measurement_concept_id"	"measurement_date"	"measurement_datetime"	"measurement_time"	"measurement_type_concept_id"	"operator_concept_id"	"value_as_number"	"value_as_concept_id"	"unit_concept_id"	"range_low"	"range_high"	"provider_id"	"visit_occurrence_id"	"visit_detail_id"	"measurement_source_value"	"measurement_source_concept_id"	"unit_source_value"	"value_source_value"

92025	702	3007876	"2002-11-10"	"2002-11-10 00:00:00"	"2002-11-10 00:00:00"	38000267	0	0	0	0	17347	1017347	"5767-9"	3007876	"{nominal}"	"Cloudy urine (finding)"

92026	702	3007876	"2002-11-10"	"2002-11-10 00:00:00"	"2002-11-10 00:00:00"	38000267	0	0	0	0	17347	1017347	"5767-9"	3007876	"{nominal}"	"Urine smell ammoniacal (finding)"

92027	702	3008204	"2002-11-10"	"2002-11-10 00:00:00"	"2002-11-10 00:00:00"	38000267	0	0	0	0	17347	1017347	"32167-9"	3008204	"{nominal}"	"Translucent (qualifier value)"

92028	702	3006513	"2002-11-10"	"2002-11-10 00:00:00"	"2002-11-10 00:00:00"	38000267	0	0.8	0	8840	0	17347	1017347	"20505-4"	3006513	"mg/dL"	"0.8"

92029	702	3023539	"2002-11-10"	"2002-11-10 00:00:00"	"2002-11-10 00:00:00"	38000267	0	7.8	0	8840	0	17347	1017347	"5797-6"	3023539	"mg/dL"	"7.8"

92030	702	3035350	"2002-11-10"	"2002-11-10 00:00:00"	"2002-11-10 00:00:00"	38000267	0	0	0	0	17347	1017347	"2514-8"	3035350	"{nominal}"	"Urine ketone test = +++ (finding)"

92031	702	3022621	"2002-11-10"	"2002-11-10 00:00:00"	"2002-11-10 00:00:00"	38000267	0	5.6	0	8482	0	17347	1017347	"5803-2"	3022621	"pH"	"5.6"

92032	702	3005897	"2002-11-10"	"2002-11-10 00:00:00"	"2002-11-10 00:00:00"	38000267	0	280.9	0	8840	0	17347	1017347	"5804-0"	3005897	"mg/dL"	"280.9"

92033	702	3021601	"2002-11-10"	"2002-11-10 00:00:00"	"2002-11-10 00:00:00"	38000267	0	0	0	0	17347	1017347	"5802-4"	3021601	"{nominal}"	"Urine nitrite negative (finding)"

92034	702	3000330	"2002-11-10"	"2002-11-10 00:00:00"	"2002-11-10 00:00:00"	38000267	0	1.0	0	0	0	17347	1017347	"5811-5"	3000330	"{nominal}"	"1.0"

observation

"observation_id"	"person_id"	"observation_concept_id"	"observation_date"	"observation_datetime"	"observation_type_concept_id"	"value_as_number"	"value_as_string"	"value_as_concept_id"	"qualifier_concept_id"	"unit_concept_id"	"provider_id"	"visit_occurrence_id"	"visit_detail_id"	"observation_source_value"	"observation_source_concept_id"	"unit_source_value"	"qualifier_source_value"

120529	38824	4060985	"1971-04-27"	"1971-04-27 00:00:00"	38000280	0	0	0	986518	1986518	"162864005"	4060985

120530	38824	4323208	"1945-02-12"	"1945-02-12 00:00:00"	38000280	0	0	0	986525	1986525	"428251008"	4323208

120531	38825	43054909	"2020-04-30"	"2020-04-30 00:00:00"	38000280	0	0	0	986531	1986531	"72166-2"	43054909

120532	38825	37311060	"2020-03-12"	"2020-03-12 00:00:00"	38000280	0	0	0	986527	1986527	"840544004"	37311060

120533	38825	4323208	"2018-03-29"	"2018-03-29 00:00:00"	38000280	0	0	0	986528	1986528	"428251008"	4323208

120534	38826	4060985	"1991-07-26"	"1991-07-26 00:00:00"	38000280	0	0	0	986536	1986536	"162864005"	4060985

120535	38826	4240902	"1952-10-29"	"1952-10-29 00:00:00"	38000280	0	0	0	986533	1986533	"91935009"	4240902

120536	38826	433644	"1952-10-29"	"1952-10-29 00:00:00"	38000280	0	0	0	986533	1986533	"300913006"	433644

120537	38826	37311060	"2020-03-07"	"2020-03-07 00:00:00"	38000280	0	0	0	986539	1986539	"840544004"	37311060

120538	38826	43054909	"2019-11-01"	"2019-11-01 00:00:00"	38000280	0	0	0	986538	1986538	"72166-2"	43054909

visit_occurrence

"visit_occurrence_id"	"person_id"	"visit_concept_id"	"visit_start_date"	"visit_start_datetime"	"visit_end_date"	"visit_end_datetime"	"visit_type_concept_id"	"provider_id"	"care_site_id"	"visit_source_value"	"visit_source_concept_id"	"admitting_source_concept_id"	"admitting_source_value"	"discharge_to_concept_id"	"discharge_to_source_value"	"preceding_visit_occurrence_id"

2310604	91491	9202	"2019-05-11"	"2019-05-11 00:00:00"	"2019-05-11"	"2019-05-11 00:00:00"	44818517	"c6a8d650-bd6c-49fc-9c71-0cd052b0d30e"	0	0	0	2310602

2310605	91491	9202	"2019-06-10"	"2019-06-10 00:00:00"	"2019-06-10"	"2019-06-10 00:00:00"	44818517	"d93cc0b7-b810-4c23-b9ca-5392c3532847"	0	0	0	2310604

2310608	91491	9203	"2019-07-22"	"2019-07-22 00:00:00"	"2019-07-22"	"2019-07-22 00:00:00"	44818517	"211c8837-6875-4e34-b613-ee8654db1a3a"	0	0	0	2310605

2310607	91491	9202	"2019-11-25"	"2019-11-25 00:00:00"	"2019-11-25"	"2019-11-25 00:00:00"	44818517	"9d56b132-a81c-42be-9fa5-f0d056c3a038"	0	0	0	2310608

2310603	91491	9202	"2020-03-04"	"2020-03-04 00:00:00"	"2020-03-04"	"2020-03-04 00:00:00"	44818517	"bd03466d-efc8-4e0d-85cd-e3d0ebc14e1c"	0	0	0	2310607

2310614	91492	9202	"2002-02-11"	"2002-02-11 00:00:00"	"2002-02-11"	"2002-02-11 00:00:00"	44818517	"fadefca1-251f-4d75-864b-6bed039a0807"	0	0	0

2310609	91492	9202	"2002-03-13"	"2002-03-13 00:00:00"	"2002-03-13"	"2002-03-13 00:00:00"	44818517	"75e86492-c21c-44da-bcb2-f9dbfc150974"	0	0	0	2310614

2310615	91492	9202	"2003-02-17"	"2003-02-17 00:00:00"	"2003-02-17"	"2003-02-17 00:00:00"	44818517	"8f7f5895-e313-46d0-91f5-c97ef5212350"	0	0	0	2310609

2310612	91492	9202	"2003-09-22"	"2003-09-22 00:00:00"	"2003-09-22"	"2003-09-22 00:00:00"	44818517	"8312fbd4-ba42-42f8-b404-8b194f9f019b"	0	0	0	2310615

2310616	91492	9202	"2004-02-23"	"2004-02-23 00:00:00"	"2004-02-23"	"2004-02-23 00:00:00"	44818517	"692ebef8-d8e8-47d5-9bce-418642128b56"	0	0	0	2310612

visit_detail

"visit_detail_id"	"person_id"	"visit_detail_concept_id"	"visit_detail_start_date"	"visit_detail_start_datetime"	"visit_detail_end_date"	"visit_detail_end_datetime"	"visit_detail_type_concept_id"	"provider_id"	"care_site_id"	"visit_detail_source_value"	"visit_detail_source_concept_id"	"admitting_source_value"	"admitting_source_concept_id"	"discharge_to_source_value"	"discharge_to_concept_id"	"preceding_visit_detail_id"	"visit_detail_parent_id"	"visit_occurrence_id"

1953351	37586	9202	"2010-06-27"	"2010-06-27 00:00:00"	"2010-06-27"	"2010-06-27 00:00:00"	44818517	"c5ee92a9-0699-464f-8d0c-5a630fad9365"	0	0	0	953351

1953347	37586	9202	"2010-07-27"	"2010-07-27 00:00:00"	"2010-07-27"	"2010-07-27 00:00:00"	44818517	"0634e1db-eff0-419f-9fd8-a887abde09bf"	0	0	0	1953351	953347

1953352	37586	9202	"2011-07-03"	"2011-07-03 00:00:00"	"2011-07-03"	"2011-07-03 00:00:00"	44818517	"20c67a2c-da8f-4fd7-a9be-a1a1447fed75"	0	0	0	1953347	953352

1953353	37586	9202	"2012-07-08"	"2012-07-08 00:00:00"	"2012-07-08"	"2012-07-08 00:00:00"	44818517	"fe073903-1b4b-4d30-82e6-5a22ee4fb842"	0	0	0	1953352	953353

1953354	37586	9202	"2013-07-14"	"2013-07-14 00:00:00"	"2013-07-14"	"2013-07-14 00:00:00"	44818517	"e8110d95-bc95-4ee3-969b-02e4e57f8cf0"	0	0	0	1953353	953354

1953355	37586	9202	"2014-07-20"	"2014-07-20 00:00:00"	"2014-07-20"	"2014-07-20 00:00:00"	44818517	"db97b5eb-3ebc-409f-888c-ea5975e7018b"	0	0	0	1953354	953355

1953356	37586	9202	"2015-07-26"	"2015-07-26 00:00:00"	"2015-07-26"	"2015-07-26 00:00:00"	44818517	"9b20f1e9-fac0-498c-8769-c31e44ed38b6"	0	0	0	1953355	953356

1953357	37586	9202	"2016-07-31"	"2016-07-31 00:00:00"	"2016-07-31"	"2016-07-31 00:00:00"	44818517	"2e7c35c9-8f24-4c42-b384-11d1e8e82068"	0	0	0	1953356	953357

1953348	37586	9202	"2017-08-06"	"2017-08-06 00:00:00"	"2017-08-06"	"2017-08-06 00:00:00"	44818517	"ec2bf47a-26ec-4747-a6c0-4496cfea0e21"	0	0	0	1953357	953348

1953358	37586	9202	"2018-08-12"	"2018-08-12 00:00:00"	"2018-08-12"	"2018-08-12 00:00:00"	44818517	"0bc9bc92-2a89-4cae-86a8-dbefbba7eea5"	0	0	0	1953348	953358

procedure_occurrence

"procedure_occurrence_id"	"person_id"	"procedure_concept_id"	"procedure_date"	"procedure_datetime"	"procedure_type_concept_id"	"modifier_concept_id"	"quantity"	"provider_id"	"visit_occurrence_id"	"visit_detail_id"	"procedure_source_value"	"procedure_source_concept_id"	"modifier_source_value"

147921	22459	4195645	"2020-05-22"	"2020-05-22 00:00:00"	38000267	0	567669	1567669	"169690007"	4061811

147922	22459	4295880	"2019-07-12"	"2019-07-12 00:00:00"	38000267	0	567670	1567670	"76601001"	4295880

147923	22459	4060626	"2020-05-22"	"2020-05-22 00:00:00"	38000267	0	567669	1567669	"169230002"	4060626

147924	22459	4168236	"2020-05-22"	"2020-05-22 00:00:00"	38000267	0	567669	1567669	"274804006"	4168236

147925	22459	4234541	"2020-05-22"	"2020-05-22 00:00:00"	38000267	0	567669	1567669	"90226004"	4234541

147926	22461	4199235	"1995-12-18"	"1995-12-18 00:00:00"	38000267	0	567687	1567687	"434158009"	4199235

147927	22461	4335825	"1996-06-15"	"1996-06-15 00:00:00"	38000267	0	567679	1567679	"433236007"	4335825

147928	22461	4326177	"1995-08-22"	"1995-08-22 00:00:00"	38000267	0	567724	1567724	"430193006"	4326177

147929	22461	4163872	"1996-06-15"	"1996-06-15 00:00:00"	38000267	0	567679	1567679	"399208008"	4163872

147930	22461	4163872	"1996-06-15"	"1996-06-15 00:00:00"	38000267	0	567681	1567681	"399208008"	4163872

death

"person_id"	"death_date"	"death_datetime"	"death_type_concept_id"	"cause_concept_id"	"cause_source_value"	"cause_source_concept_id"

33389	"2020-03-05"	"2020-03-05 00:00:00"	38003566	37311061	"840539006"	37311061

67097	"2014-09-06"	"2014-09-06 00:00:00"	38003566	381316	"230690007"	381316

75672	"2011-09-05"	"2011-09-05 00:00:00"	38003566	321042	"410429000"	321042

75531	"2012-12-20"	"2012-12-20 00:00:00"	38003566	258780	"185086009"	258780

46235	"2016-09-07"	"2016-09-07 00:00:00"	38003566	321042	"410429000"	321042

90319	"1980-01-04"	"1980-01-04 00:00:00"	38003566	4129902	"126906006"	4129902

58930	"2020-03-20"	"2020-03-20 00:00:00"	38003566	37311061	"840539006"	37311061

73226	"1995-01-12"	"1995-01-12 00:00:00"	38003566	378419	"26929004"	378419

91592	"1966-12-23"	"1966-12-23 00:00:00"	38003566	79740	"109838007"	79740

74381	"1946-06-06"	"1946-06-06 00:00:00"	38003566	4329847	"22298006"	4329847

concept

"concept_id"	"concept_name"	"domain_id"	"vocabulary_id"	"concept_class_id"	"standard_concept"	"concept_code"	"valid_start_date"	"valid_end_date"	"invalid_reason"

2727412	"Bypass Right Brachial Artery to Right Lower Arm Artery with Autologous Venous Tissue, Open Approach"	"Procedure"	"ICD10PCS"	"ICD10PCS"	"S"	"0317093"	"2014-04-22"	"2099-12-31"

2727413	"Bypass Right Brachial Artery to Upper Arm Vein with Autologous Venous Tissue, Open Approach"	"Procedure"	"ICD10PCS"	"ICD10PCS"	"S"	"031709D"	"2014-04-22"	"2099-12-31"

2727414	"Bypass Right Brachial Artery to Lower Arm Vein with Autologous Venous Tissue, Open Approach"	"Procedure"	"ICD10PCS"	"ICD10PCS"	"S"	"031709F"	"2014-04-22"	"2099-12-31"

43015547	"Bypass Right Brachial Artery to Superior Vena Cava with Autologous Venous Tissue, Open Approach"	"Procedure"	"ICD10PCS"	"ICD10PCS"	"S"	"031709V"	"2017-05-14"	"2099-12-31"

1524540	"Bypass Right Brachial Artery to Lower Extremity Vein with Autologous Venous Tissue, Open Approach"	"Procedure"	"ICD10PCS"	"ICD10PCS"	"S"	"031709W"	"2020-01-01"	"2099-12-31"

2888298	"Medical and Surgical @ Upper Arteries @ Bypass @ Brachial Artery, Right @ Open @ Autologous Arterial Tissue"	"Procedure"	"ICD10PCS"	"ICD10PCS Hierarchy"	"S"	"03170A"	"1970-01-01"	"2099-12-31"

2727415	"Bypass Right Brachial Artery to Right Upper Arm Artery with Autologous Arterial Tissue, Open Approach"	"Procedure"	"ICD10PCS"	"ICD10PCS"	"S"	"03170A0"	"2014-04-22"	"2099-12-31"

2727416	"Bypass Right Brachial Artery to Right Lower Arm Artery with Autologous Arterial Tissue, Open Approach"	"Procedure"	"ICD10PCS"	"ICD10PCS"	"S"	"03170A3"	"2014-04-22"	"2099-12-31"

2727417	"Bypass Right Brachial Artery to Upper Arm Vein with Autologous Arterial Tissue, Open Approach"	"Procedure"	"ICD10PCS"	"ICD10PCS"	"S"	"03170AD"	"2014-04-22"	"2099-12-31"

2727418	"Bypass Right Brachial Artery to Lower Arm Vein with Autologous Arterial Tissue, Open Approach"	"Procedure"	"ICD10PCS"	"ICD10PCS"	"S"	"03170AF"	"2014-04-22"	"2099-12-31"

concept_relationship

"concept_id_1"	"concept_id_2"	"relationship_id"	"valid_start_date"	"valid_end_date"	"invalid_reason"

46210207	46210207	"Mapped from"	"2018-07-31"	"2099-12-31"

46210207	46210207	"Maps to"	"2018-07-31"	"2099-12-31"

46210209	46067302	"Has AMP"	"2015-01-31"	"2099-12-31"

46210209	46210209	"Mapped from"	"2018-07-31"	"2099-12-31"

46210209	46210209	"Maps to"	"2018-07-31"	"2099-12-31"

46210211	46210211	"Mapped from"	"2018-07-31"	"2099-12-31"

46210211	46210211	"Maps to"	"2018-07-31"	"2099-12-31"

46210212	46067305	"Has AMP"	"2015-01-31"	"2099-12-31"

46210212	46210212	"Mapped from"	"2018-07-31"	"2099-12-31"

46210212	46210212	"Maps to"	"2018-07-31"	"2099-12-31"

concept_ancestor

"ancestor_concept_id"	"descendant_concept_id"	"min_levels_of_separation"	"max_levels_of_separation"

19091998	40911987	2	2

19101013	2026458	1	1

19091901	35857675	0	0

19086233	2073947	2	2

19085606	43208045	1	1

19083433	43143118	2	2

19081141	41160098	2	2

19078781	36066671	1	2

19063926	36271128	1	2

19075394	21150086	1	1

vocabulary

"vocabulary_id"	"vocabulary_name"	"vocabulary_reference"	"vocabulary_version"	"vocabulary_concept_id"

"Death Type"	"OMOP Death Type"	"OMOP generated"	"44819135"

"Cohort Type"	"OMOP Cohort Type"	"OMOP generated"	"44819234"

"Condition Status"	"OMOP Condition Status"	"OMOP generated"	"32887"

"NUCC"	"National Uniform Claim Committee Health Care Provider Taxonomy Code Set (NUCC)"	"http://www.nucc.org/index.php?option=com_content&view=article&id=107&Itemid=132"	"2018-06-26 NUCC"	"44819137"

"Revenue Code"	"UB04/CMS1450 Revenue Codes (CMS)"	"http://www.mpca.net/?page=ERC_finance"	"2010 Release"	"44819133"

"ICD9CM"	"International Classification of Diseases, Ninth Revision, Clinical Modification, Volume 1 and 2 (NCHS)"	"http://www.cms.gov/Medicare/Coding/ICD9ProviderDiagnosticCodes/codes.html"	"ICD9CM v32 master descriptions"	"5046"

"Type Concept"	"OMOP Type Concept"	"OMOP generated"	"Type Concept 20210212"	"32808"

"Race"	"Race and Ethnicity Code Set (USBC)"	"http://www.cdc.gov/nchs/data/dvs/Race_Ethnicity_CodeSet.pdf"	"Version 1.0"	"44819109"

"Domain"	"OMOP Domain"	"OMOP generated"	"44819147"

"CMS Place of Service"	"Place of Service Codes for Professional Claims (CMS)"	"http://www.cms.gov/Medicare/Medicare-Fee-for-Service-Payment/PhysicianFeeSched/downloads//Website_POS_database.pdf"	"2009-01-11"	"44819110"

relationship

"relationship_id"	"relationship_name"	"is_hierarchical"	"defines_ancestry"	"reverse_relationship_id"	"relationship_concept_id"

"Has specimen proc"	"Has specimen procedure (SNOMED)"	false	false	"Specimen proc of"	44818775

"Has specimen source"	"Has specimen source identity (SNOMED)"	false	false	"Specimen identity of"	44818776

"Has specimen morph"	"Has specimen source morphology (SNOMED)"	false	false	"Specimen morph of"	44818777

"Has specimen topo"	"Has specimen source topography (SNOMED)"	false	false	"Specimen topo of"	44818778

"Has specimen subst"	"Has specimen substance (SNOMED)"	false	false	"Specimen subst of"	44818779

"Has due to"	"Has due to (SNOMED)"	false	false	"Due to of"	44818780

"Has relat context"	"Has subject relationship context (SNOMED)"	false	false	"Relat context of"	44818781

"Has dose form"	"Has dose form (SNOMED)"	false	false	"Dose form of"	44818782

"Occurs after"	"Occurs after (SNOMED)"	false	false	"Occurs before"	44818783

"Has asso proc"	"Has associated procedure (SNOMED)"	false	false	"Asso proc of"	44818784

**Recall heatmaps**

Full-context outputs achieved recall close to 1.0 across nearly all models in both datasets, confirming that F1 gains at higher context levels were driven primarily by improved recovery of eMERGE-positive patients. In limited-context synthetic settings, recall was highly variable and often near zero, consistent with the overly restrictive query logic described in the main text. In the UNC EHR setting, limited-context recall was more variable than near-zero, but full-context Level 3 outputs were more uniformly high. The persistent PPV limitation rather than recall deficit explains the F1 ceiling in real-world data.

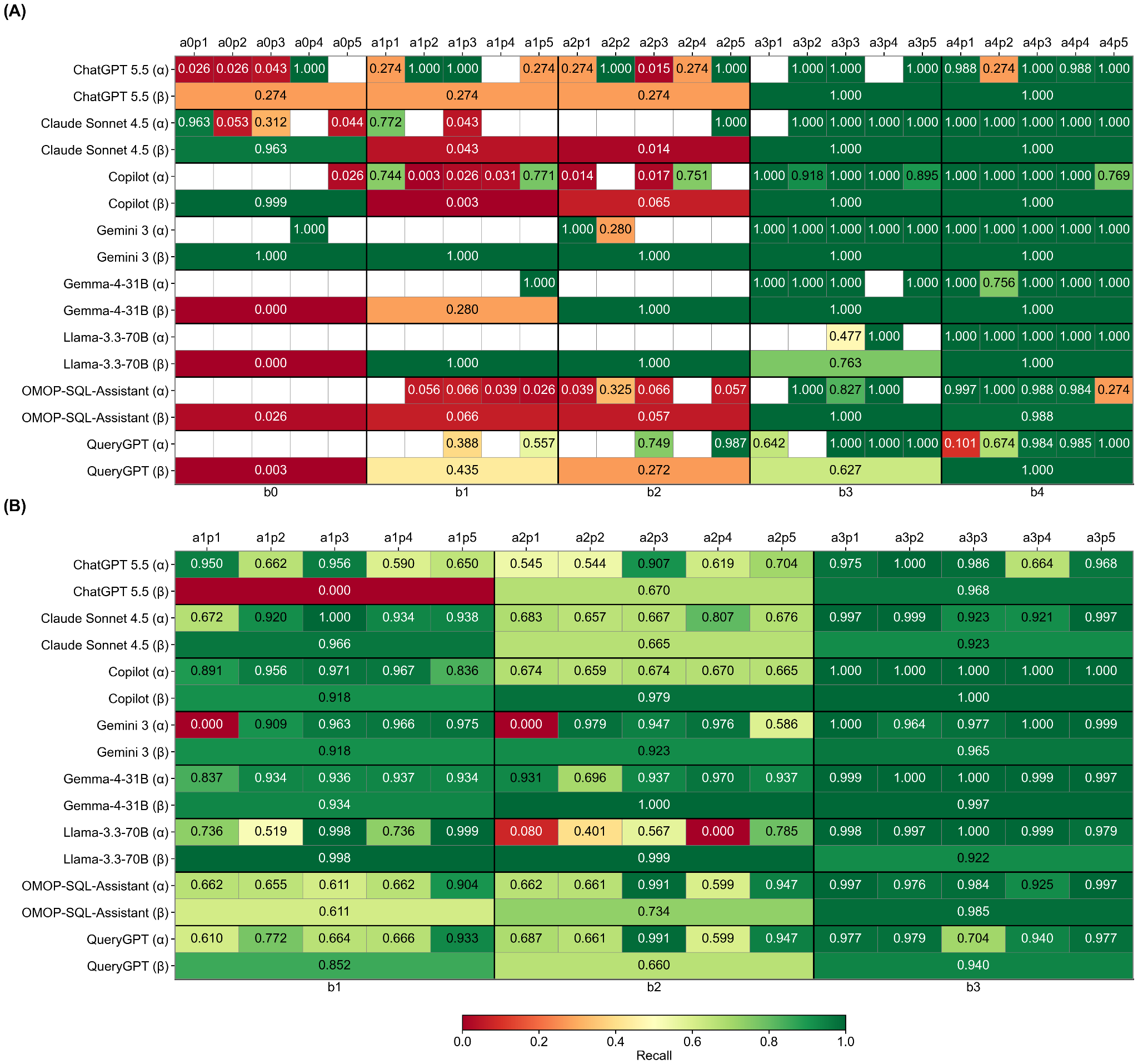

**Figure S1. Recall heatmap by model and prompt in (A) synthetic and (B) UNC EHR data.** Rows and columns follow the same structure as Figure 1. Full-context outputs commonly achieved high recall, indicating improved recovery of eMERGE-positive patients.

Alt text: (A) Synthetic OMOP recall heatmap showing highly variable or non-evaluable results at lower context levels, with generally near-perfect recall at Levels 3–4. (B) UNC EHR recall heatmap showing variable performance at Levels 1–2 and consistently high recall at Level 3.
